# Improving HIV Pre Exposure Prophylaxis (PrEP) uptake and initiation: process evaluation and recommendation development from a national PrEP programme

**DOI:** 10.1101/2022.10.09.22280871

**Authors:** Claudia S Estcourt, Jen MacDonald, John Saunders, Rak Nandwani, Ingrid Young, Jamie Frankis, Dan Clutterbuck, Nicola Steedman, Lisa McDaid, Jenny Dalrymple, Paul Flowers

## Abstract

**Background:** HIV pre-exposure prophylaxis (PrEP) is key to HIV transmission elimination but implementation is challenging and under-researched. We undertook a process evaluation of the first two years of a national PrEP programme to explore barriers and facilitators to implementation and to develop recommendations to improve implementation, focussing on PrEP uptake and initiation.

**Methods:** Stage 1 involved semi-structured telephone interviews and focus groups (09/2018-07/2019) with geographically and demographically diverse patients seeking/using/declining/stopping PrEP (n=39), sexual healthcare professionals (n= 54), community-based organisation service users (n=9) and staff (n=15) across Scotland. We used deductive thematic analysis, to derive and then map key barriers and facilitators to priority areas that experts agreed would enhance initiation and uptake. In Stage 2 we used analytic tools from implementation science to systematically generate evidence-based, theoretically-informed recommendations to enhance uptake and initiation of PrEP.

**Results:** Barriers and facilitators were multi-levelled and interdependent. Barriers included the rapid pace of implementation without additional resource, and a lack of familiarity with PrEP prescribing. Facilitators included opportunities for acquisition of practice-based knowledge and normalisation of initiation activities. We refined our 68 “long-list” recommendations to 41 using expert input and the APEASE criteria. Examples include: provision of PrEP in diverse settings to reach all in need; co-produced, culturally sensitive training resources for healthcare professionals, with focused content on non-daily dosing; meaningful collaborative working across all stakeholders.

**Conclusions:** These evidence-based, theory informed recommendations provide a robust framework for optimising PrEP uptake and initiation in diverse settings to ensure PrEP reaches all who may benefit.

**Summary for table of contents:** Zero new HIV infections could become a reality if HIV pre-exposure prophylaxis (PrEP) programmes are successfully implemented but the World Health Organisation recognizes that large scale roll out is challenging.

We used implementation science research tools in novel ways to evaluate one of the world’s first national PrEP programmes, to develop evidence-based recommendations for use across a range of settings to improve PrEP uptake and initiation.

Adopting these recommendations could enable governments and societies to better address HIV prevention goals.

## Background

HIV pre-exposure prophylaxis (PrEP), in which people take antiretroviral medication to prevent HIV acquisition, is a major advance in biomedical prevention of HIV. In clinical trials, orally administered PrEP has been shown to reduce the risk of HIV acquisition by 44-97% (1-4). Although PrEP is becoming increasingly available, insights from real-world implementation studies are limited (5-7). The World Health Organization and others acknowledge the importance of making PrEP available for safe, effective prevention outside clinical trial settings as key to realising its potential to end HIV epidemics (8,9). Implementation science tools could help unlock the full potential of PrEP (10) to assist with the elimination of HIV transmission (9).

Scotland became one of the first countries worldwide to implement a national PrEP programme (11). At the time, there were around 4600 people living with HIV attending specialist care in Scotland (12) and 228 people newly diagnosed with HIV each year, half of whom were gay, bisexual, and other men who have sex with men (GBMSM) (13). From July 2017, PrEP and all associated medical monitoring were made available free at point of access, as part of broader HIV combination prevention and sexual health care, almost exclusively through sexual health clinics, to those at greatest risk of HIV acquisition (14). Prescribing followed specialist association guidance (15), but services developed their own local models of delivery, largely within existing budgets. These broadly involved: [1] identifying a patient as a PrEP candidate; [2] provision of PrEP information, baseline screening for HIV and other blood borne viruses (BBVs), sexually transmitted infections (STIs), and renal function; [3] prescribing and dispensing PrEP; and [4] regular in person reviews for HIV, BBV, and STI testing, renal monitoring, adherence support, wider sexual health promotion, and PrEP prescribing (15). Quantitative outcomes from the national PrEP Programme have been reported as part of routine surveillance (12-14) and through detailed epidemiology (6).

We conducted a process evaluation of the first two years of Scotland’s PrEP programme. Our approach divided the PrEP care cascade into three sections; awareness and access (16), initiation and uptake and adherence and retention in care (17). Here we focussed on uptake and initiation of PrEP.

We addressed the following research questions:

1. Within PrEP care pathways where exactly should we intervene (priority areas) to optimise uptake and initiation?
2. What are the barriers and facilitators to optimising implementation within these priority areas?
3. Which evidence-based and theoretically informed recommendations could improve the implementation of PrEP uptake and initiation?

## Methods

As described elsewhere (16-17), Stage 1 is a retrospective qualitative process evaluation within a larger natural experimental design study evaluating PrEP implementation in Scotland (research questions 1 and 2), and Stage 2 involves development of recommendations to improve PrEP uptake and initiation, using systematic intervention development approaches (research question 3).

### Data collection

#### Participants

We used multi-perspective purposive sampling to understand the implementation of PrEP uptake and initiation from diverse viewpoints. In total, 117 participants took part in individual semi-structured telephone interviews (n=71) or in one of 10 group discussions (n=46) (September 2018-July 2019). The sample comprised: 39 patients; 54 healthcare professionals; nine non-governmental organisation (NGO) service users; and 15 NGO staff from across Scotland. All NGOs had an HIV prevention remit and served GBMSM, trans, and/or Black African communities. Group discussions included one type of stakeholder only.

Patients were either using PrEP (n=23, 59%) or had declined (n=5, 13%), stopped (n=6, 15%), or been assessed as ineligible (n=5, 13%) for PrEP. PrEP users included those who took PrEP daily, event-based or both ways. They ranged in age from 20-72 years with just over half (n=21, 54%) between 25-34 years. All self-identified as gay or bisexual men, the majority of whom (n=34, 87%) were cisgender. Almost all were of ‘White British’ (n=31, 80%) or ‘Other White’ (n=7, 18%) ethnicity. Two thirds had a university degree (n=26, 67%) and the majority were in employment (n=34, 87%). The patient areas of residence reflected a mix of relative affluence and deprivation although the most (n=5, 16.7%) and least (n=3, 10%) deprived quintiles (according to Scottish Index of Multiple Deprivation (SIMD), which divides areas into five subgroups according to the extent to which an area is “deprived” (18)) were under-represented and patients predominantly resided in the middle three quintiles (73%) (data missing for 9 participants). Healthcare professionals were all involved in PrEP implementation in a mix of rural (n=12, 22%), semi-rural/urban (n=8, 15%), or urban (n=34, 63%) settings, largely reflecting the wider Scottish population distribution. They included specialist sexual health doctors and nurses of various grades, some with national PrEP roles, PrEP prescribing general practitioners (who prescribed PrEP on the Scottish islands), health promotion officers, a midwife, and a clinical secretary responsible for PrEP-related administration. NGO service users were all of Black African ethnicity, predominantly cis-gender women, and not using PrEP.

#### Recruitment

Healthcare professionals offered patients the opportunity to take part in the study during routine consultations taking place in four of the 14 regional health boards (responsible for the protection and improvement of their population’s health) providing over 90% of PrEP related care in Scotland. NGO service users who were either engaged with NGOs *and* attending sexual health clinics (classed as patients above) or only engaged with NGO services (classed as NGO service users above) were invited to participate via interactions with NGO staff. We recruited these and other NGO staff and healthcare professionals across all of Scotland’s 14 regional health boards by email invitation.

#### Procedure

All participants provided informed verbal or written consent immediately prior to the interviews /group discussions. We collected data with the aid of a topic guide that included open-ended questions designed to explore participants’ experiences and perceptions of uptake and initiation of PrEP, rather than questions based on any theoretical concepts anticipated to influence implementation. Where possible within the group discussions, dialogue between participants was encouraged rather than between facilitators and participants. All participants talked from their own and others’ perspectives; data were taken at face value. Patients were offered a £30 shopping voucher as reimbursement for their time.

Data collection was led by JM, with input from experienced qualitative researchers, PF, IY, and JF. JM, PF, IY, and JF reviewed and discussed early transcripts for quality assurance purposes. All interviews and group discussions were audio recorded, transcribed verbatim, anonymised, and imported into NVivo software for analysis.

### Data analysis

#### Stage 1

##### Research Question 1: Within PrEP care pathways where exactly should we intervene (priority areas) to optimise uptake and initiation?

Firstly, we used the Action, Actor, Context, Target, Time framework (19), to conceptualise the sequential actors, actions, settings, and processes that constituted PrEP adherence and retention in care. Secondly, we iteratively created a series of visualisations of the overall behavioural system of PrEP adherence and retention in care using available UK guidance on best clinical practice in PrEP provision (12) and transcripts of early interviews and group discussions. Thirdly, we comprehensively assessed the breadth and depth of data relating to the patient pathway through PrEP adherence and retention in care. Finally, we (PF & JM) ranked the most important areas which were considered to be amenable to change to create priority areas for intervention. Then research team members with real-world clinical experience of providing PrEP services in assorted settings (CSE, RN, JS) provided further input resulting in the identification of nine final priority areas for recommendation development.

##### Research Question 2: What are the barriers and facilitators to implementing the priority areas for PrEP adherence and retention in care?

We (JM and PF) conducted deductive thematic analysis (20) of the qualitative data concerning barriers and facilitators for each priority area. We used the relative frequency of barriers and facilitators to manage the volume of findings and to ensure we focussed only on those that were deemed most important. This stage ended with the identification of the major barriers and facilitators for the priority areas.

#### Stage 2

##### Research question 3: Which evidence-based and theoretically informed recommendations could improve PrEP adherence and retention in care?

We treated each of the priority areas independently and analysed each separately. Firstly, we entered the key barriers and facilitators into a matrix. Secondly, we used the Behaviour Change Wheel (BCW) approach (21), and systematically coded the key barriers and facilitators for each priority area using the Theoretical Domains Framework (TDF) (22). Finally, we specified corresponding Intervention Functions (broad ways of intervening relevant to the theoretical domains) and used the Behaviour Change Technique (BCT) and corresponding Taxonomy (BCTTV1v1) (23) to describe, in detail and using a standardised language, potential intervention content that may be helpful to operationalise the Intervention Functions, address key barriers and facilitators, and enhance future PrEP implementation. This created an initial “long-list” of recommendations. All coding and drafting of recommendations were completed by JM and double-checked for accuracy, validity, and credibility by PF. Any disagreements were discussed until consensus was reached.

Finally, clinical expert team members (CE, RN, JS) scrutinised, sense-checked, and shortlisted the long list of initial recommendations using the APEASE criteria (24). This resulted in the introduction of some new recommendations, in addition to minor amendments to or merging/deleting of existing recommendations.

### Ethical considerations

The Glasgow Caledonian University Research Ethics Committee (HLS/NCH/17/037, HLS/NCH/17/038, HLS/NCH/17/044) and the South East Scotland National Health Service Research Ethics Committee (18/SS/0075, R&D GN18HS368) provided ethical approval.

## Results

### Research Question 1: Within PrEP care pathways where exactly should we intervene (priority areas) to optimise uptake and initiation?

Nine priority areas for intervention (black) were identified from the wider range of potential areas of focus (Figure 1). Each potential area forms part of a typical patient pathway at the start of PrEP care. The priority areas involve two actors (sexual healthcare professionals (HCPs) and potential PrEP users (patients)).

**Figure 1:**
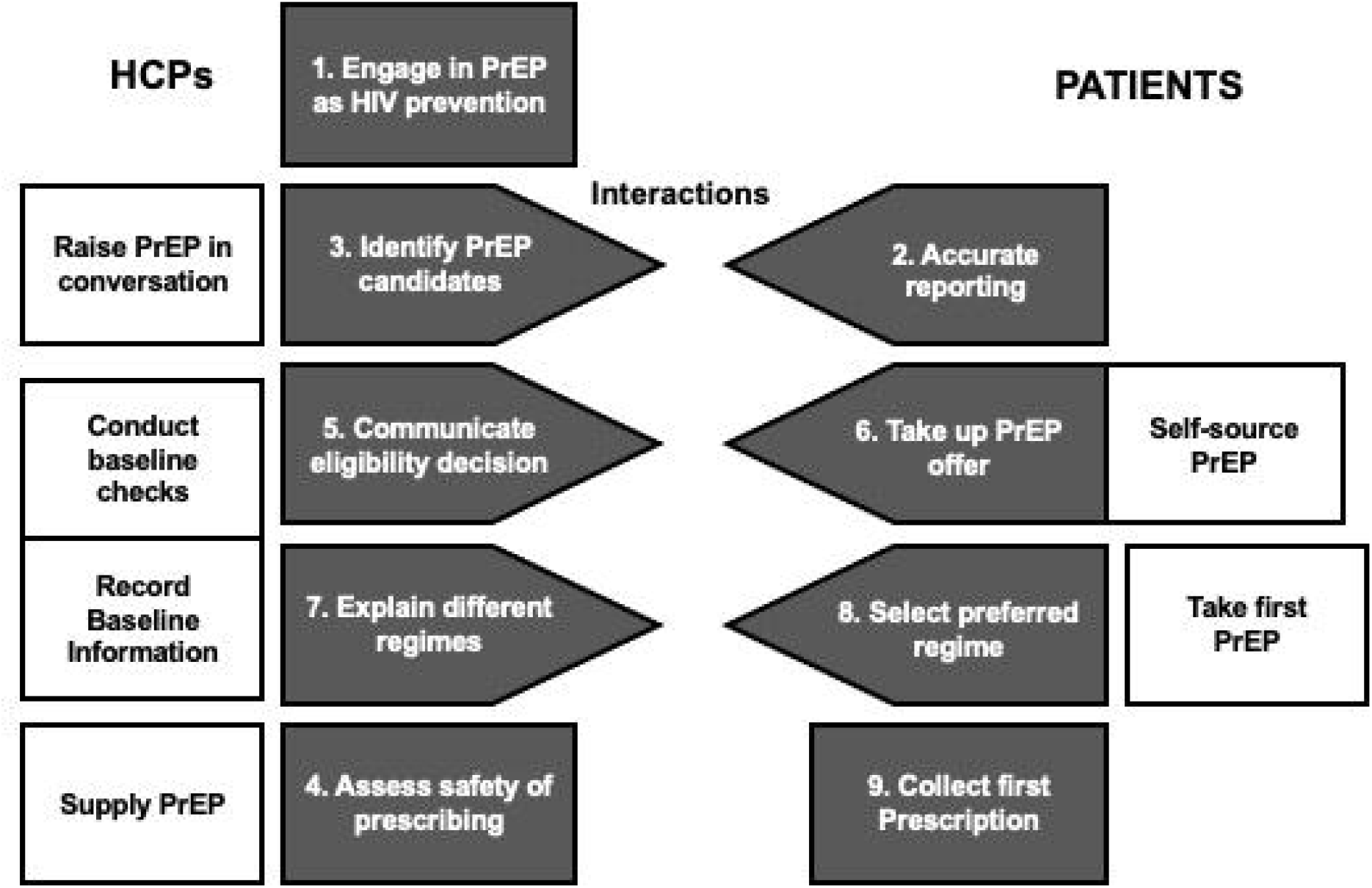
Steps in the uptake and initiation of PrEP illustrating where to intervene to improve implementation. Shaded boxes depict areas for recommendation development. (1) HCPs engaging with PrEP as an acceptable approach to HIV prevention; (2) Potential PrEP users accurately reporting HIV risk behaviour; (3) HCPs identifying PrEP candidates based on risk of HIV acquisition; (4) HCPs determining safety of prescribing and medical suitability for PrEP; (5) HCPs communicating eligibility/ineligibility for PrEP; (6) Potential PrEP users taking up PrEP; (7) HCPs adequately explaining different PrEP regimens; (8) Potential PrEP users choosing their preferred regimen; and (9) Potential PrEP users obtaining their first PrEP prescription. Steps in clear boxes were not selected as priority areas. Pointed Boxes highlight the interactions between the steps. Connected boxes highlight the associated nature of those steps.

### Research Question 2: What were the barriers and facilitators to optimising implementation within these priority areas?

In general, facilitators to implementing the priority areas in one service directly matched corresponding barriers in others (Table 1). Even before systematically generating recommendations, the analysis began to directly highlight useful lessons learned about implementation.

**Table 1:** The major barriers and facilitators to each of the nine priority areas within uptake and initiation of PrEP.

| Agreed priority area for intervention (i.e. recommendation development) | Key barriers | Key facilitators |
| --- | --- | --- |
| **1) HCPs engage with PrEP as an approach to HIV prevention** | - lack of dedicated budget, pace of implementation and competing service innovations (e.g. HPV vaccination of GBMSM)<br>- beliefs about being de-skilled by PrEP initiation due to its repetitive nature<br>- moral views on PrEP, condom use, STIs and homophobic attitudes | -collegiality, team work, and peer-support fostered formal and informal networks and relationships at multiple levels.<br>-enhanced job role and job satisfaction associated with PrEP initiation reinforced the work<br>-staff understood the bigger picture and understood the efficacy and cost-effectiveness of PrEP relative to care costs associated with people living with HIV.<br>-staff had insight into the social and emotional consequences of HIV and PrEP for the individual<br>-staff recognized the role PrEP has in bringing people whose behaviours |
|  |  | and/or behaviours of others put them at highest risk of HIV to specialist services |
| **(2) Potential PrEP users accurately report their HIV risk behaviour** | -patient concerns over meeting eligibility criteria confounds accurate reporting<br>-patient expectations of being judged by HCPs constrains accurate reporting<br>-low levels of sexual, sexual health and HIV literacy make frank conversations about HIV risk very hard | -the very availability of PrEP enables worthwhile frank conversations about actual HIV risks<br>-expectations that HCPs will be approachable, culturally sensitive and non-judgmental |
| **(3) HCPs identify PrEP candidates based on risk of HIV acquisition** | -difficulties operationalising eligibility criteria<br>-there were doubts concerning veracity of patient accounts of their HIV risks (e.g. inflating their reported risk to meet eligibility criteria) | -they could build on prior expertise around HIV risks particularly amongst GBMSM<br>-peer support and discussions about eligibility are useful and added new skills<br>-longstanding competencies in communication skills around sexual/drug histories could be employed<br>-beliefs that PrEP can enable open and honest disclosures of HIV risk behaviours<br>-supportive IT systems and documentation enable identification of PrEP candidates |
| **(4) HCPs determine safety of prescribing and medical suitability for PrEP** | -HCPs worried about making the wrong decisions around prescribing and some believed that PrEP prescribing should be consultant (specialist medic)-led<br>-there were limited opportunities to take up education and training<br>-conflicting advice and mixed messages from senior colleagues made the situation unclear | -HCPs felt comfortable with prescribing given their previous experience with post exposure prophylaxis (PEP) and HIV care<br>- formal and informal training and learning opportunities at local-, regional-, and national-levels were available<br>-formal and informal opportunities for peer support were available (e.g., to |
|  | -prescribing PrEP was sporadic and not routine | seek advice, check and share decision-making, and discuss more medically complex cases, at local-, regional-, and national-levels)<br>-frequent opportunities to prescribe PrEP and on the job experience<br>-booked PrEP appointments provide the opportunity to prepare for interactions by reviewing electronic patient records |
| **(5) HCPs communicate eligibility/ineligibility for PrEP** | -they felt under pressure from patients to provide PrEP<br>-they lacked knowledge, skills and experience to convey risk/benefits of PrEP effectively | -they could make explicit reference to the eligibility criteria to shape their decisions<br>-they could discuss ineligibility in a positive light and use it as a teachable moment for wider HIV risk reduction<br>-they could suggest self-sourcing PrEP online and the offer of monitoring within the SHS as an alternative to free NHS prescription<br>-they can focus on risk/benefits for given individuals |
| **(6) Potential PrEP users take up offer of PrEP** | -they are reticent to take daily medication<br>-they are put-off by the perceived health and social consequences (e.g., side effects and perceived potential reputational damage)<br>-HCP are perceived to push PrEP<br>-they are dubious about the effectiveness of PrEP | -they can tailor regimes flexibly (i.e., daily and or event based)<br>-they want to take PrEP because of the perceived health and social consequences (e.g., HIV risks and better sex)<br>-PrEP use is reinforced by significant others (peers, partners, friends)<br>-HCPs provide a balanced narrative and enable informed tailored choices around PrEP |
|  |  | -they are confident in the efficacy of PrEP |
| (7) HCPs explain the different PrEP regimens | -they lack familiarity with on-demand dosing | -they can use information booklets and illustrations to show how to follow on-demand dosing to structure conversations |
| (8) Potential PrEP users choose their preferred regimen | -HCPs offer limited dosing regimens not suited to patients' life circumstances | -HCPs offer a range of appropriate regimen choices in a balanced manner<br>-there is considerable information of PrEP dosing available on-line |
| (9) Potential PrEP users get their first PrEP prescription | -there are delays to starting PrEP whilst waiting for baseline HIV test results<br>-PrEP is only available through off-site dispensing | -there is on-site dispensing |

Here we provide a brief narrative overviewing the details in Table 1 for each priority area (1-9) along with indicative quotations from participants for context.

#### [1] Engaging HCPs with PrEP as an HIV prevention approach

Whilst structural issues related to capacity within the sector, “We’re having to squeeze this extra work into the same resource.” (HCP), psychosocial issues encompassed factors such as staff attitudes. Facilitators included collegiality, peer-fostered support, and the use of existing networks to actively share innovation:

> ‘*We were all able to share things like protocols, and how we were all working…so that nurses will be able to prescribe. These are all things that are being worked on together, so that each health board doesn’t need to do things individually, and I think that helped hugely’* (HCP).

#### [2] PrEP users accurately reporting their own HIV risk behaviour and/or other factors placing them at higher risk of HIV acquisition

Several psychosocial issues were identified including the importance of sexual and sexual health literacy and expectations of staff being approachable and non-judgmental:

> “*There’s a moral judgement that comes with clinical risk assessment, and patients can pick up on that, and they pick up on it really, really quickly, and that just wrecks a patient’s consultation*.*”* (HCP)
>
> “*It’s a question of just listening a little bit more. Not having a dismissive attitude. I think everybody likes to be listened to. And it’s really important, when people, even if they are speaking with an accent, to try and listen, and try to understand where they are coming from*” (CBO staff working with Black African communities)

#### [3] HCPs correctly identifying PrEP candidates

HCPs were comfortable raising PrEP with GBMSM but experienced difficulties with women and some minoritised groups. This was partly because HCPs felt that the PrEP eligibility criteria (ref HPS yr 1 report) aligned with question areas they would not necessarily ask non-GBMSM.

However, supportive IT systems, which highlighted eligibility criteria were felt to facilitate PrEP conversations:

> *‘‘Through years of experience. I make it [assessing patient’s HIV risk] so matter of fact as if it’s conversation and I think a lot of my colleagues do the same*.*”* (HCP)

#### [4] HCP determining the safety of prescribing

Issues such as familiarity with HIV medication, training and peer support were important:

> *It’s definitely a learning process. Experience, really, and the more exposure to it [PrEP] has definitely changed the way that I think, and assess people. And what the follow-up is as well*.*”* (HCP)

#### [5] Communicating eligibility decisions

Knowledge, skills and experience were key.

> *“I think that terminology makes patients really angry. And I think that is probably one of the biggest problems, is telling people, you’re ‘not eligible’. I think that people really don’t like being told that” (HCP)*
>
> *“It’s not that you’re making that decision, so I would sit with the guidelines and go through them one by one with like the criteria, and go through them and say ‘you don’t fit any of them’*.*” (HCP)*

#### [6] Patients taking up the offer of PrEP

The way HCP present choices around PrEP was important, as were the beliefs of others (e.g., peers, partners) and PrEP users’ own beliefs about PrEP efficacy and the perceived consequences of PrEP.

> *“I think her words were, have you thought about PrEP? She [doctor] sort of prompted it, prompted the conversation but didn’t push it and then I continued the conversation*.*” (PrEP user)*
>
> *“He [clinic nurse] was kind of telling me about all the good things about PrEP, but I wasn’t*…*I don’t know. I didn’t want to buy it, if this is a phrase, because he was almost saying that it’s the best thing ever, because he was using it, he was using it and he told me that. So, I don’t know, I kind of stopped using the [clinic]*.*” (PrEP user)*

#### [7] HCPs adequately explaining the different PrEP regimens

Some staff struggled because of their lack of experience with on-demand dosing in particular.

> *“I don’t know how good I would be if they were saying so I’m going to have sex on a Saturday and then I’m going to have sex on a Thursday, when do I actually start and stop it, you know? So, it’s case-by-case and I probably still need to refresh my memory a little bit and read up a bit on that still if I was doing that because most of the people are just taking it every day*.*” (HCP)*

#### [8] Potential PrEP users choosing their preferred regimen

The importance of choosing a dosing regimen that was tailored to their life circumstances was felt to be key.

#### [9] Potential PrEP users getting their first prescription. The practicalities of where PrEP was dispensed were particularly important

> *“It [hospital pharmacy] is not the easiest place to get to if you don’t have your own transport*.*” (HCP)*

### Research Question 3: Which evidence-based and theoretically informed recommendations should improve future PrEP uptake and initiation?

Analysis of the main barriers and facilitators to each priority area enabled us to systematically theorise what was working well in relation to implementation, and also what was not. We were then able to formulate specific tailored recommendations to enhance the future implementation of each of the priority areas in both general terms (intervention functions) and highly specific terms (operationalised BCTTV1s) (Table 2). Full details of our underpinning analysis are provided within supplementary files.

**Table 2:** Specific recommendations to improve the implementation of uptake and initiation using the behaviour change wheel approach, incorporating the behaviour change technique taxonomy.

| **Agreed priority area for intervention (i.e. recommendation development)** | **Key recommendations to enhance the implementation of uptake and initiation**<br>(Numbers in brackets relate to the BCT from the BCTTV1) |
| --- | --- |
| **1) HCPs engage with PrEP as an approach to HIV prevention** | 1.1 Ensure those that fund sexual health services provide the resource to match the costs of the programme<br>1.2 Ensure a realistic timescale for PrEP implementation that allows for critical planning activities, such as estimating the likely demand for PrEP, conducting a full service review to determine capacity and how PrEP will fit into existing practices, and working in partnership across the whole HIV sector to develop and deliver an 'official' national PrEP training package (9.1), including examples of how to deliver PrEP services (4.1, 6.1), to prepare the workforce (12.1, 12.2). Such training should also focus on enhancing the cultural competencies of all staff to work with diverse communities (4.1, 6.1, 8.1, 2.2)<br>1.3 Ensure a multileveled national infrastructure has a clear remit to promote, |
|  | coordinate, and monitor HCP engagement with PrEP (12.2, 2.1)<br>1.4 In the early stages of PrEP roll-out, national PrEP coordination groups and local PrEP leaders should organise shared learning events and ensure formal and informal peer support systems are in place (e.g. real-time/email support from senior staff, team meetings, 'phone a friend', clinical network arrangements) to strengthen working relationships among HCPs (12.2, 3.1, 3.2, 6.2)<br>1.5 Use local, regional, and national infrastructures to foster a team-oriented, 'open-source' approach to PrEP-related work (e.g. share protocols, training materials, service innovations and adaptations, insights into how to engage HCPs with PrEP) (12.2, 3.1, 3.2, 6.1, 6.2)<br>1.6 Identify HCPs with a strong belief in and commitment to PrEP to act as local champions and inspire and engage other HCPs with PrEP (12.2)<br>1.7 Educate HCPs on the economic and wider benefits and value of PrEP for the healthcare system, local sexual health services, communities, and individual clients, for example, by informing of the positive health, cost/ financial, service engagement, social, and emotional impacts of PrEP (e.g. talks from leading clinicians in favour of PrEP, positive testimonials of PrEP users) (5.1, 5.3, 5.6, 9.1) |
| **(2) Potential PrEP users accurately report their HIV risk behaviour** | 2.1 Sexual health services could ask CBO staff who have high levels of cultural competency in delivering sexual health promotion interventions to Black Africans, trans people, and cis women to share their tailored vocabularies and co-produce a stock of key phrases and scenarios to enable HCPs to sensitively probe clients when taking a sexual/ drug history (4.1, 6.1, 7.1)<br>2.2 Ensure HCPs are educated (5.1), trained (4.1, 6.1, 8.1, 8.7), and appraised in their skills (2.2) in explaining the risk-benefit of PrEP and mandate this activity in a formal protocol (4.1, 5.1)<br>2.3 Ensure PrEP information and communications (e.g. sexual health service and CBO staff-client interactions, national patient information booklets, sexual health service, CBO, and HIV/PrEP activists' websites and social media, marketing campaigns) avoid using the term 'eligibility criteria' and instead adopt 'needs-based' terminology that explicitly conveys the risks and benefits of PrEP (5.1, 13.2)<br>2.4 HCPs should actively promote PrEP to clients as one of several sexual health promotion methods (5.1) and emphasise their own and other experts and credible sources' support for it (e.g. government, public health agencies, CBO staff) (9.1) |
|  | 2.5 Facilitate and maintain (e.g. via training, clinical supervision, reflective practice) a warm, welcoming, and friendly atmosphere wherein HCPs communicate with clients in a non-judgemental manner, using active listening and inclusive, sex- and PrEP-positive, and destigmatising language to establish trust and ensure an open dialogue (12.2, 5.3) |
| **(3) HCPs identify PrEP candidates based on risk of HIV acquisition** | 3.1 Ensure PrEP information and communications (e.g. sexual health service and CBO staff-client interactions, national patient information booklets, sexual health service, CBO, and HIV/PrEP activists' websites and social media, marketing campaigns) avoid using the term 'eligibility criteria' and instead adopt 'needs-based' terminology that explicitly conveys the risks and benefits of PrEP (5.1, 13.2)<br>3.2 Adopt a protocolled approach to PrEP that includes advice (e.g. clear statements and nuanced examples) regarding the eligibility criteria (4.1, 13.2)<br>3.3 Ensure HCPs maintain their knowledge of the HIV risks among different groups, and skills in conducting culturally sensitive clinical risk assessments (e.g. ongoing professional development, clinical supervision) (5.1, 2.2, 2.3, 8.1)<br>3.4 Ensure a range of peer-support systems are in place (e.g. real-time/email support, team meetings, 'phone a friend', clinical network arrangements) to assist HCPs in making complex eligibility decisions (12.2, 3.1, 3.2, 6.2)<br>3.5 HCPs should actively but sensitively promote PrEP to clients as a method for HIV prevention (5.1) and emphasise their own and other experts and credible sources' support for it (e.g. government, public health agencies, CBO staff) (9.1) so clients feel comfortable to disclose their HIV risks |
| **(4) HCPs determine safety of prescribing and medical suitability for PrEP** | 4.1 Produce national guidelines to promote and instruct HCPs on safe prescribing of and medical suitability for PrEP, review and update the guidelines to reflect new information and lessons learned over time (5.1, 4.1)<br>4.2 Use national infrastructure to facilitate discussion among senior clinicians and reach a consensus on best practice for a range of scenarios to promote consistency in decisions on the safety of prescribing and medical suitability for PrEP (12.2, 3.1, 3.2)<br>4.3 Ensure HCPs are educated about PrEP via a comprehensive and ongoing training package that covers HIV testing, the HIV window period, and risk of antiretroviral resistance, common side-effects and their typically transient nature, the likelihood of toxic effects and role of monitoring to prevent long-term issues, and |
|  | contraindications (5.1)<br>4.4 Ensure there are formal and informal peer-support systems at local-, regional-, and national-level (e.g. real-time/email support, team meetings, 'phone a friend', clinical network arrangements) to assist HCPs in making complex decisions on medical suitability for PrEP (12.2, 3.1, 3.2, 6.2)<br>4.5 Demystify PrEP and build HCPs confidence by presenting PrEP as a drug that can be prescribed by any qualified prescriber or supplied via agreed protocols (e.g. PGD) within sexual health service settings (13.2)<br>4.6 National coordinated PrEP training should include inter-disciplinary online PrEP learning resources for HCPs which can be broken down into short modules on specific topics (e.g. covering safe prescribing of and medical suitability for PrEP) and spread out over a period of time (5.1, 4.1). These could be aligned with professional development for many job roles (12.2)<br>4.7 Introduce a shadowing scheme across different sexual health services to enable HCPs from services with few PrEP users to become familiar with PrEP processes, including ensuring safe prescribing of and medical suitability for PrEP (12.2, 6.1)<br>4.8 Train HCPs on how to conduct adequate assessments of any underlying health conditions and interpret the results of new tests required to establish medical suitability for PrEP (4.1, 6.1), share example cases for HCPs to discuss and work through (8.1, 8.7), provide feedback (2.2), and allow opportunities for ongoing reflections on skill acquisition (2.3)<br>4.9 Inform HCPs that they can easily access up-to-date and evidence-based online information on interactions between PrEP and other drugs (e.g. www.hiv-druginteractions.org) (4.1) |
| **(5) HCPs communicate eligibility/ineligibility for PrEP** | 5.1 Adopt a protocolled approach to PrEP that includes advice (e.g. clear statements and nuanced examples) regarding the eligibility criteria (4.1, 13.2)<br>5.2 Throughout PrEP provision and promotion (e.g. during HCP and CBO staff-client interactions, in national patient information booklets, on sexual health service, CBO, and HIV/ PrEP activists' websites and social media, in marketing campaigns) avoid using the term 'eligibility criteria' and instead adopt 'needs-based' terminology that explicitly conveys PrEP decisions as a function of the individual risk-benefit of PrEP for each client (12.2, 13.2) |
|  | 5.3 Ensure HCPs are educated, trained, and appraised in their skills in discussing the risks and benefits of PrEP (e.g. through online modules, peer support, clinical supervision), for example, by giving information on PrEP health consequences (5.1), producing a ‘how to’ script for common PrEP scenarios based on the lessons learned of SHCPs with general medicine expertise (4.1, 7.1), and providing opportunities to shadow (6.1), practice with (8.1, 8.7), and receive feedback (2.2) from more experienced HCPs<br>5.4 HCPs should reassure clients that they are at low risk for HIV by educating them (e.g. verbally, directing to reputable websites) on the facts of HIV transmission and effectiveness of alternative sexual health promotion methods (5.1)<br>5.5 HCPs need to be aware of the option to self-source PrEP and could consider directing clients who do not meet the eligibility criteria but would still like to access PrEP to reputable online sources of information about where to buy PrEP (e.g. provision of national patient information booklets, signpost to appropriate websites (3.1)<br>5.6 HCPs should explore the root cause(s) of HIV-related anxieties among clients who do not have an identified need for PrEP and work with them to problem solve solutions (1.2) |
| **(6) Potential PrEP users take up of PrEP** | 6.1 All sectors involved in PrEP should consider a range of approaches (e.g. via HCP-/CBO-client interactions, sexual health service, CBO, and HIV/PrEP activists’ websites and social media, national patient information booklets, marketing campaigns) to: normalise PrEP by drawing parallels to the use of daily preventive medicine in other areas of health (e.g. contraceptive pill to protect against pregnancy, blood thinners to reduce the risk of heart attack and stroke) (13.2); and educate potential PrEP users on the flexibility of PrEP by informing them of the idea of ‘seasons of risk’ (i.e. unlikely to be on PrEP forever, can start and stop as circumstances dictate) and the various dosing options (i.e. can opt for less intensive on-demand dosing, if appropriate) (5.1, 13.2)<br>6.2 HCPs should draw on research evidence and what they know about other patients’ decision-making and experiences to inform patients of the health, social, and emotional benefits of PrEP (5.1, 5.3, 5.6, 16.3) but also stress that PrEP is a choice and discuss the pros and cons of taking up PrEP compared to not taking up PrEP with respect to clients’ individual interests (9.2)<br>6.3 HCPs should educate clients about the potential side-effects of PrEP and their |
|  | typically transient nature (5.1), share management strategies for the most common side-effects (1.2), and reassure against concerns about longer-term toxic effects by drawing attention to the tests undertaken at regular reviews (5.1)<br>6.4 Co-produced PrEP information and communications (e.g. HCP-/CBO staff-client interactions, national patient information booklets, SHS, CBO, and HIV/PrEP websites and social media, posters in sexual health service and CBO settings, marketing campaigns) should provide an accessible, scientific explanation of what PrEP does (i.e. how it works inside the body) and describe PrEP efficacy and safety with reference to key research and 'real world' studies and regional or national HIV incidence data (5.1, 9.1) |
| **(7) HCPs explain the different PrEP regimens** | 7.1 Use a variety of ways to educate HCPs about on-demand dosing (4.1) and assist them during consultations (7.1). For example:<br>• Develop a range of resources (e.g. brief fact sheet, PrEP provider pocket guide, national patient information booklets) with clear written instructions and diagrams that depict how to take PrEP on-demand, including examples of when to start and stop for various scenarios, which can be used to educate HCPs (4.1) and assist them during consultations (7.1). Such resources should ideally be co-produced by a range of diverse organisations and the communities who will use them)<br>• Provide HCPs with laminated copies of the on-demand dosing diagrams that they can pin to their wall as a quick reminder of how to take PrEP on-demand (4.1, 7.1)<br>• Record a short video or soundbite that explains on-demand dosing for different scenarios that HCPs may watch or listen to at a future date (4.1)<br>• Include an online or paper-based quiz with questions about on-demand dosing as part of HCPs PrEP training and ongoing professional development and ensure that there is opportunity to discuss answers (2.7) |
| **(8) Potential PrEP users choose their preferred regimen** | 8.1 HCPs should inform clients of their options for how to take PrEP by way of a balanced narrative (5.1) and then jointly, with each individual client, facilitate a decisional balance weighing up the pros and cons per option, taking into account lifestyle and/or the availability of evidence to support it (i.e. dependent on gender and whether oral, anal, or vaginal/frontal sex) (9.2)<br>8.2 HCPs and CBO staff could direct clients to reputable online sources of information on the various ways to take PrEP (e.g. sexual health service, CBO, and |
|  | HIV/PrEP activists' websites and social media) (3.1, 9.1) in addition to the information they provide (e.g. verbally, via provision of national patient information booklet) |
| **(9) Potential PrEP users get their first PrEP prescription** | 9.1 Ensure services establish a PrEP supply chain (12.2) and maintaining agreed stock levels (12.5) to enable HCPs to dispense PrEP as soon as possible<br>9.2 Work with pharmacy leads to extend the role of community pharmacists to enable clients to obtain PrEP via a range of settings (12.1) |

**Legend:** Full details of our underpinning analysis are provided within supplementary files. Details of the operationalisation of behaviour change techniques are shown in brackets.

## Discussion

Complex multi-levelled factors shaped PrEP implementation. Nine specific areas of the PrEP care cascade involved in initiation and uptake of PrEP were both amenable to change and prioritised for improvement. The corresponding barriers and facilitators were multi-levelled and interdependent. Many were psychosocial, relating directly to the way staff or patients thought and felt; others related to the organisation of services, wider issues of access to support and training, and factors relating to the environmental infra-structure of services. Using tools from implementation science, we systematically generated highly specific, theoretically informed and evidence-based ways of optimising PrEP implementation in the future. Examples include: provision of PrEP in diverse settings to reach all in need; co-produced, culturally sensitive training resources for healthcare professionals, with focused content on non-daily dosing; meaningful collaborative working across all stakeholders.

To date, several attempts have been made to conceptualise the implementation of PrEP but these have been largely broad and descriptive, typically categorising the whole of PrEP care into four or five large steps within a continuous, linear ‘care cascade’ (25-28). Published studies have tended to focus on using these high-level steps to audit or quantify PrEP implementation, seeking to identify and understand key points of attrition within particular populations and associated health care systems (29). There are numerous examples of PrEP prescribing guidance (15,30-31), but fewer published studies specifically address the implementation of PrEP routine care pathways and services. A scoping review of PrEP delivery models (32) created a comprehensive inventory of existing models, but did not specifically focus on delivery of the detailed steps of the PrEP cascade within the models described. A review of PrEP implementation identified multiple barriers to PrEP uptake, some of which mirrored those we described (33). The authors proposed multilevel interventions to target these barriers but acknowledge that proposed interventions do not always align to specific barriers.

In contrast, no work to date has used conceptualisations of the care cascade as a starting point for systematic, focussed service improvement whilst explicitly using theory and evidence to enhance implementation. We directly addressed this gap by taking a single key step of the PrEP care cascade, the ‘uptake and initiation of PrEP’, and focussed on it as an area in need of intervention development to enhance future implementation. We derived recommendations (interventions) directly from the barriers and facilitators at each priority area.

Some recommendations warrant additional comment. In relation to ‘engaging HCPs with PrEP as an acceptable approach to HIV prevention’, we highlight the need to address both structural *and* psychosocial issues. We also emphasise the importance of considering financial and other resources as well as the timescale for implementation. These factors are likely to be central to HCP engagement which in turn is central to patient uptake. We also recommend a multileveled national infrastructure to promote, coordinate, and monitor HCP engagement with PrEP and highlight how these structural initiatives could be bolstered by a range of local initiatives such as engaging staff through local “PrEP champions”. The barriers these recommendations are designed to overcome were strikingly similar to those reported in a number of studies within Pinto et al’s recent review (33).

In relation to ‘potential PrEP users accurately reporting their HIV risk behaviour…”, we found that depending on the cultural context, it may be important to educate and persuade HCP about the ‘bigger picture’ of PrEP provision and overcome any residual moralism and stigma relating to sex, homophobia, or racism which has also been described in other studies (33-34). Stigma is well recognised as a potent barrier to accessing HIV testing, prevention and care and it also might inhibit the full disclosure of HIV acquisition risk factors such as stigmatised sexual behaviours or partner numbers relevant to PrEP offer and uptake. Stigma may also apply to and inhibit the taking of PrEP itself (35-37). We recommend close partnership work between sexual health services, CBOs and PrEP users to enable sensitive, culturally appropriate conversations around PrEP, and to help HCPs improve their cultural competencies. The strongly supported health care and community-level “PrEP-positive” ethos described by our participants seems highly appropriate and would need to be extended to all settings in which PrEP may be provided in the future, particularly those in which sexual health is less familiar.

Our findings suggest that the ‘PrEP eligibility criteria’ which were used by HCPs to help identify people who might benefit most from PrEP (26), should be reframed and understood as needs-based approaches to HIV prevention, conveying the pros and cons of PrEP so that it can be extended to all who could benefit. This could largely remove the issue that criteria are less sensitive for identifying people from certain groups or racial backgrounds as also reported in other countries (38).

A large epidemiological analysis published after this study showed that Scottish implementation models strongly favour GBMSM and have limited reach into other key vulnerable populations (6,14). In parallel, the characteristics of people newly diagnosed with HIV in Scotland have changed since the introduction of PrEP and now people are more likely to have acquired HIV though heterosexual sex and to be non-white indigenous than in the pre-PrEP era (14,39), similar to findings from Australia (40). As noted in our recommendations and by others, reaching all groups that could benefit from PrEP is essential (9); Several studies provide explanations for low PrEP uptake in some key vulnerable populations. Among women of colour in the UK, important factors were low awareness of PrEP, feelings of stigma related to HIV itself and attending sexual health clinics, and a preference for trusted community settings for discussion about HIV testing and prevention (35,41)). Among people who inject drugs in Scotland, awareness of PrEP was low but some would find PrEP appealing if provided within familiar settings such as outreach drug services (42). Very few trans people have accessed PrEP in Scotland (12). International studies suggest that the need for PrEP among this group is high but important barriers to access preclude uptake (36,43). Restricting PrEP provision to sexual health clinics probably deters some trans people who could benefit (44). Additional or tailored recommendations to enhance PrEP uptake and initiation for people from vulnerable populations are needed as evidence accrues.

We used a novel, rigorous approach to developing recommendations which is not typical of approaches to enhancing implementation. The resulting recommendations are anchored in the evidence and theory-driven (22) and are specified using a standardised language to describe intervention content in detail (i.e., intervention functions and behaviour change techniques (23)). Typically, the initial stages of the PrEP care cascade involve a complex patient journey, marked by setting-specific interactional dynamics and a series of interdependent joint and individual behaviours. Our adoption of a behavioural lens, and the subsequent systematic development of highly specific ways to enhance implementation, meant we re-conceptualised this patient journey as a series of distinct and sequential behaviours. This approach led to costs and benefits; where we gained through this behavioural specificity and our ability to specify future recommendations in great detail (e.g. behavioural change techniques), there was duplication of effort to detail shared antecedents to the varied behaviours.

We focussed on one national context and although findings are likely to be generalisable to similar settings, it is uncertain how recommendations might apply in very different contexts. In particular, as all PrEP care was free of charge, participants did not face the financial barriers reported from some settings (45). Very few people in Scotland on PrEP are not GBMSM (13) and our findings lack specificity for other groups. A high proportion of PrEP user participants had a university qualification and while representative of those on PrEP in Scotland, the sample under-represents those with lower health and PrEP literacy who may have other needs and preferences for accessing PrEP care. Furthermore, the COVID-19 pandemic led to a reconfiguration of some sexual health and PrEP services and our findings may be more or less relevant as a result. Our evaluation took place relatively early in the PrEP programme which probably magnifies early stage issues which become less important as familiarity increases.

To support individuals and populations to fully benefit from PrEP we must overcome the considerable challenges of large-scale implementation (31). Here, we combined qualitative data from multiple viewpoints and used multiple analytic tools to systematically detail useful insights concerning initiation and uptake from the first two years of Scottish PrEP implementation. To our knowledge, we present the first evidence-based and theory-informed recommendations which can be used flexibly across a range of settings to improve PrEP initiation and uptake. Our findings will inform future Scottish implementation of PrEP (46) and could usefully contribute to the global public health priority of elimination of HIV transmission by 2030 (31,47).

## Data Availability

Due to the sensitive nature of the questions asked in this study, survey respondents were assured raw data would remain confidential and would not be shared.

## Declarations

### Ethics approval and consent to participate

The study received ethical approval from the Glasgow Caledonian University Research Ethics Committee (REC) (HLS/NCH/17/037, HLS/NCH/17/038, HLS/NCH/17/044) and the South East Scotland NHS REC (18/SS/0075, R&D GN18HS368)

### Consent for publication

Not applicable

### Authors’ contributions

All authors contributed to the conception and design of the studies, interpretation of findings, revision of the manuscript and approved the final version. Specific additional contributions are as follows and marked where appropriate in the manuscript: CSE was principal investigator and involved in all stages of the research and wrote the initial draft of the manuscript. PF conceptualised the design of the process evaluation and led the behavioural analyses. JM led the study day to day and undertook all research activities including data collection and analysis under the supervision of PF and CSE. JS, RN, DC, NS and CSE provided expert clinical interpretation. IY and JF contributed to data collection and analysis. JD led the ethical approval application.

## Acknowledgements

We are very grateful to the users, patients and staff of sexual health services in all 14 Health Boards, Drs Ruth Holman, Dan Clutterbuck, Maggie Gurney, Nil Banerjee, Pauline McGough, Daniela Brawley, Kirsty Abu-Rajab, Hame Lata, Anne McLellan, Alison Currie, Sharon Cameron, Hilary MacPherson, Janice Irvine, Graham Leslie, Ciara Cunningham, Maggie Watts. We thank staff and users of HIV Scotland; Waverley Care (SX Project and African Health Project); THT Scotland; Hwupenyu Health and Wellbeing; and Scottish Trans Alliance. We thank Nathan Sparling and Jacqueline Gray for their contributions to the research process.

