## Appendix 1 for "Improving HIV Pre Exposure Prophylaxis (PrEP) uptake and initiation: process evaluation and recommendation development from a national PrEP programme"

| **Appendix 1. The TDF and BCW analysis underpinning the evidence-based and theoretically informed recommendations** | | | | | | | | |
| --- | --- | --- | --- | --- | --- | --- | --- | --- |
| **Priority areas for intervention** | **Barriers** | **Facilitators** | **Indicative quotes** | **TDF domains** | **Intervention functions** | **Potential BCTs** | **Recommendations for those considering implementing PrEP at scale** | **Decision re: kill/keep/modify with notes** |
| ***PrEP provision*** | | | | | | | | |
| SHCPs engage with PrEP as an approach to HIV prevention | SHCPs find it difficult to engage with PrEP as an approach to HIV prevention because of frustrations about how the PrEP programme was implemented (e.g. quickly, within existing budgets, coinciding with the introduction of the HPV vaccination programme for MSM, without staff consultation, with limited training opportunities) and displacement of other services | -- | “*M1: When PrEP was rolled out last year, if it had come with a dozen extra staff…*  *F4: Great training programme, lots of support.*  *F2: Yeah.*  *M1: …to do all these extra clinics, which have happened, then, and the rest of the service had gone on as it had done before, I think we'd all be smiling and happy, and would think this was great. So the issue is, we’re having to squeeze this extra work into the same resource…*  *F4: A service that’s been already squeezed*.” (SHCPs) | Environmental context and resources  Social influences | Enablement  Environmental restructuring  Persuasion  Training | 3.2 Social support (practical)  5.1 Information about health consequences  5.3 Information about social and environmental consequences  9.1 Credible source  12.1 Restructure the physical environment  12.2 Restructure the social environment  12.5 Adding objects to the environment  4.1 Instruction on how to perform the behaviour  6.1 Demonstration of the behaviour  8.1 Behavioural practice/rehearsal  2.2 Feedback on behaviour | 1. Those that fund SHS should provide the resource required to match the innovation (i.e. increase the budget according to predicted PrEP demand to ensure adequate capacity for effective implementation and scale-up) in the initial months of national rollout (3.2). A business case produced by senior HIV clinicians that also outlines the health benefits of PrEP (5.1) and potential future savings of PrEP implementation within the healthcare system (i.e. more cost-effective than spending on HIV treatment) (5.3) could be helpful in this regard (9.1)  2. Government and public health agencies should ensure that the roll-out of PrEP does not coincide with the introduction of other programmes (12.1, 12.2) or if this is unavoidable / it is preferable to make a major change through introducing two innovations at once (i.e. so one period of disruption not two), that appropriate resources are devoted to measured service reorganisation (3.2)  3. Government and public health agencies responsible for PrEP should ensure a well-paced timescale for PrEP implementation that allows for critical planning activities, such as estimating the likely demand for PrEP, conducting a full service review to determine capacity and how PrEP will fit into existing practices, and working in partnership across the whole HIV sector to develop and deliver an ‘official’ national PrEP training package (9.1), including examples of how to deliver PrEP services (4.1, 6.1), to prepare the workforce (12.1, 12.2). Such training should also focus on enhancing the cultural competencies of all staff to work with diverse communities (4.1, 6.1, 8.1, 2.2)  50. Ensure that there are mechanisms (e.g. email, huddles, team meetings) in place to keep SHCPs informed about PrEP implementation and involve them in proposed service reorganisation by providing formal (e.g. consultations) and informal (e.g. suggestion box) opportunities to share any concerns and feedback ideas for improvements (12.2, 12.5)  51. Identify SHCPs with a strong belief in and commitment to PrEP to act as local champions and inspire and engage other SHCPs with PrEP (12.2) | 1. Keep but modify (discussed) – really important to flag that resources are needed for implementation so as not to engender animosity towards the PrEP programme.  Replace ‘innovation’ with ‘clinical activity’ or ‘costs of the programme’. Delete ‘produced by senior HIV clinicians’. *  2. Kill (no discussion) – too general and would not wish to stall PrEP just because there are other things to do.  3. Keep but modify (no discussion) – really important to flag that much more time is needed for implementation so as not to engender animosity towards the PrEP programme. Consider replacing ‘well-paced’ with ‘realistic’ and broadening HIV sector to sexual health / health sector. *  50. Kill (no discussion).  51. Keep (no discussion). |
|  | -- | SHCPs find it easy to engage with PrEP as an approach to HIV prevention because of collegiality, team work, and peer-support fostered from strong pre-existing formal and informal networks and relationships at the local-, regional- and national-level | “*There’s already existing set ups for things like HIV Lead Clinicians, Sexual Health Lead Clinicians, the Lead Nurses Services, Lead Health Promotion, these already exist, these groups of people. And so, we were all able to share things like protocols, and how we were all working, and the same goes with things like PGDs for medication, so that nurses will be able to prescribe, these are all things that are being worked on together, so that each health board doesn’t need to do things individually, and I think that helped hugely*.” (SHCP) | Environmental context and resources  Social influences | Environmental restructuring  Enablement  Modelling | 12.2 Restructure the social environment  2.1 Monitoring of behaviour by others without feedback  3.1 Social support (unspecified)  3.2 Social support (practical)  6.1 Demonstration of the behaviour  6.2 Social comparison | 4. Ensure a multileveled national infrastructure has a clear remit to promote, coordinate, and monitor SHCP engagement with PrEP (12.2, 2.1)    5. Use local, regional, and national infrastructures to foster a team-oriented, ‘open-source’ approach to PrEP-related work (e.g. share protocols, training materials, service innovations and adaptations, insights into how to engage SHCPs with PrEP) (12.2, 3.1, 3.2, 6.1, 6.2)  52. Work with SHCPs in SHS which display good multi-disciplinary relationships to understand what facilitates their team working environment and use these lessons learned to demonstrate effective team working across the region / country (6.1)  6. In the early stages of PrEP roll-out, national PrEP coordination groups and local PrEP leaders should organise shared learning events and ensure formal and informal peer support systems are in place (e.g. real-time/email support from senior staff, team meetings, ‘phone a friend’, clinical network arrangements) to strengthen working relationships among SHCPs (12.2, 3.1, 3.2, 6.2) | 4. Keep (discussed) – national coordination very important. Cannot be over emphasised.  5. Keep (no discussion) – sharing of information is key.  52. Kill (no discussion) – not specific to PrEP.  6. Keep (no discussion) – could be merged with 5. |
|  | SHCPs find it difficult to engage with PrEP as an approach to HIV prevention because of moral views on PrEP, condom use, and STIs, which are often tied up in homophobic rhetoric and unaddressed by more clinically-focused training | SHCPs find it easy to engage with PrEP as an approach to HIV prevention because they understand it’s efficacy and cost-effectiveness relative to treating HIV, have insight to the social and emotional consequences of HIV and PrEP for the individual, and recognise the role of PrEP in bringing in those at highest risk for HIV to SHS | **Example 1**  “*It doesn't talk about these conversations we're having with staff where they're saying, oh, it's going to increase STIs. All it does is tell you what PrEP is, who would benefit from it, why it's important. So it talks about the practical aspects of PrEP. It doesn't allow for a discussion around your feelings around PrEP, which I think in itself is the barrier*.” (CBO staff working with MSM)  **Example 2**  “*If you can get less diagnosis of HIV that’s a big positive. You know, in the long-term that costs more money and, you know, in terms of follow-up care, the other drugs that they would need to take, and, also, there’s the stigma if someone gets HIV. You know, I know stigma...it just hasn’t reduced. You know, I see lots of my...the clients who are HIV positive and the stigma has been a difficult thing for most of the guys*.” (SHCP) | Environmental context and resources  Behavioural regulation  Beliefs about consequences  Knowledge  Professional role and identity | Environmental restructuring  Education  Persuasion | 12.2 Restructure the social environment  5.1 Information about health consequences  5.3 Information about social and environmental consequences  5.6 Information about emotional consequences  9.1 Credible source  2.3 Self-monitoring of behaviour  12.1 Restructure the physical environment  2.7 Feedback on outcome(s) of behaviour  13.2 Framing/ reframing  13.3 Incompatible beliefs | 53. Establish and actively maintain a positive organisational culture (12.2) by educating SHCPs in a wholistic understanding of sexual health and wellbeing, equalities, heterosexism, and homophobia (5.3, 5.6, 5.1), reflecting a wholistic approach in the SHS values and mission statement and including as a core competency for professional conduct, and providing opportunities for regular reflective practice on mindfully not stigmatising groups or individuals (2.3)  26. Educate SHCPs on the economic and wider benefits and value of PrEP for the healthcare system, local SHS, communities, and individual clients, for example, by informing of the positive health, cost/ financial, service engagement, social, and emotional impacts of PrEP (e.g. talks from leading clinicians in favour of PrEP, positive testimonials of PrEP users) (5.1, 5.3, 5.6, 9.1)  27. Devise a system to monitor and evaluate the PrEP programme (12.1) and provide SHCPs with regular updates on PrEP uptake, characteristics of PrEP seekers/users, STI and HIV rates, and cost-effectiveness (e.g. via published reports at national-level, sharing of local data at team meetings) (2.7)  28. Ensure discussions on PrEP attitudes are incorporated into all PrEP training for SHCPs and that PrEP is normalised by drawing comparisons to the contraceptive pill (13.2)  29. Draw attention to SHCPs moralism of PrEP and MSM sexual behaviour and their self-identification with the principles and values of the SHS and/or a relevant professional body (e.g. as part of PrEP training, in reflective practice) (13.3) | 53. Keep but modify (discussed) – very general, not specific to PrEP. Agree with principle but not inclusive enough (e.g. to black / trans people, what about intersectionalities). However, want to ensure we retain elements such as providing a non-judgemental, stigma-free, and supportive environment.  26. Keep (no discussion).  27. Keep (no discussion). *  28. Keep but modify (discussed) – insert ‘e.g.’ before ‘by drawing comparisons to the contraceptive pill’ and expand on what we mean by this.  29. Kill (discussed) –moralistic views are more the exception than the rule. This may be relevant in less liberal countries. |
|  | SHCPs find it difficult to engage with PrEP as an approach to HIV prevention because they now have very little variation in their job role and feel that they are getting deskilled | SHCPs find it easy to engage with PrEP as an approach to HIV prevention because PrEP is enhancing their job role and satisfaction | “*F1: I opted to do [clinic], thinking that that would tick a box, keep my competencies up, keep me interested. Now we’re doing PrEP and I'm bored rigid. A monkey could do it, that’s how I feel, a monkey could do it.*  *M1: Yes, which is the unfortunate thing, because there are some complex patients there.*  *F1: Yeah, you get the odd one.*  *M1: But there's lots of very routine, just routine screens, and handing out pills*.” (SHCPs) | Environmental context and resources  Professional role and identity  Beliefs about consequences | Environmental restructuring  Persuasion | 12.2 Restructure the social environment  5.3 Information about social and environmental consequences  5.6 Information about emotional consequences | 11. Within SHS with high numbers of clients starting PrEP and/or initiating PrEP via specific clinics, rotate SHCPs to allow a range of clinical experiences (12.2)  12. SHS should explore and provide innovative ways of scheduling appointments with built-in flexibility to enable SHCPs to engage clients in discussions on wider sexual health issues (i.e. not just giving out PrEP) (12.2)  30. Ensure that all SHCPs have opportunities to keep up and develop their clinical skills (e.g. via CPD, in clinical supervision) (12.2)  31. Share positive SHCP testimonials of the ways in which PrEP is enhancing their job role and satisfaction (e.g. PrEP is a CPD opportunity, brings a more positive slant to conversations on sex and HIV risk, enables relationships with clients through continuity of care) (5.3, 5.6) | 11. Kill (no discussion) – service detail, not specific to PrEP.  12. Kill (no discussion) – not PrEP specific, applies to all health services to deliver holistic care and health improvement.  30. Kill (no discussion) – Not PrEP specific, has to happen anyway.  31. Kill (no discussion). |
| SHCPs identify PrEP candidates based on risk of HIV acquisition | SHCPs find it difficult to identify PrEP candidates based on risk of HIV acquisition because of uncertainty about the veracity of clients’ accounts of their HIV risks | SHCPs find it easy to identify PrEP candidates based on risk of HIV acquisition because the availability of PrEP allows for a more worthwhile discussion on HIV risks and thus prompts more open and honest disclosures from clients | **Example 1**  *“I don't think it's something that you can police… because you're either feeling that folk are sitting and they're thinking, they're sitting in front of you telling you what you want to hear, to fit the eligibility. Or like they’ve changed their practice, in order to get it.”* (SHCP)  **Example 2**  *“Before PrEP came on the market a lot of these guys would be telling us that they were using condoms because they thought that’s what we wanted to hear and now they’re working out that perhaps they can be a bit more honest. So, I think there’s probably a lot more honest consultations going on now.”* (SHCP) | Knowledge  Beliefs about consequences  Social influences | Education  Persuasion  Training  Enablement | 5.1 Information about health consequences  13.2 Framing/ reframing  4.1 Instruction on how to perform the behaviour  6.1 Demonstration of the behaviour  8.1 Behavioural practice/rehearsal  8.7 Graded tasks  2.2 Feedback on behaviour  9.1 Credible source | 67. Ensure PrEP information and communications (e.g. SHS-and CBO staff-client interactions, national patient information booklets, SHS, CBO, and HIV/PrEP activists’ websites and social media, marketing campaigns) avoid using the term ‘eligibility criteria’ and instead adopt ‘needs-based’ terminology that explicitly conveys the risks and benefits of PrEP and thus the rationale against its blanket supply (5.1, 13.2)  32. Ensure SHCPs are educated (5.1), trained (4.1, 6.1, 8.1, 8.7), and appraised in their skills (2.2) in explaining the risk-benefit of PrEP and mandate this activity in a formal protocol (4.1, 5.1)  68. SHCPs should explain to clients the importance of reporting an accurate sexual/ drug history to ensure they receive the most appropriate care to their individual needs (5.1, 9.1)  69. SHCPs should actively promote PrEP to clients as a method for HIV prevention (5.1) and emphasise their own and other experts and credible sources’ support for it (e.g. government, public health agencies, CBO staff) (9.1) so clients feel comfortable to disclose their HIV risks | 67. Keep but modify (not discussed) – consider deleting ‘and thus the rationale against its blanket supply’.  32. Keep (no discussion).  68. Kill (no discussion).  69. Keep but modify (discussed) – remove the section after 9.1. |
|  | SHCPs find it difficult to identify PrEP candidates based on risk of HIV acquisition because they are unsure how to navigate the ‘equivalent risk’ eligibility criterion and fear that they might stigmatise or offend non-MSM clients by asking questions to assess PrEP candidacy | SHCPs find it easy to identify PrEP candidates based on risk of HIV acquisition because they are primed to assess and identify HIV risks among MSM clients (e.g. they expect MSM to be the main group accessing PrEP, view all MSM as potentially ‘at-risk’, have a clear sense of MSM HIV risks, and are used to talking to MSM about this) | **Example 1**  *“Women who are at risk of HIV are probably pretty difficult to identify. I'd say, particularly people who are in a relationship, they’re very difficult to identify. Particularly if they don't know that someone they’re having sex with has HIV. People from minority groups, they're quite difficult to identify. But we also don't know who we don't know about, at the moment.”* (SHCP)  **Example 2**  *“We’re really well trained to know. If someone [an MSM] is telling you they’re not using condoms for anal sex or they’ve had a few burst condoms, or they’ve split up with someone and they’re having a bit of a wild three months and they’ve been quite enjoying it, and this is something they think they might want to do for a bit longer. I think everyone’s really confident at knowing straightway if someone [an MSM] would benefit from PrEP.”* (SHCP) | Environmental context and resources  Knowledge  Beliefs about consequences  Skills | Enablement  Education  Training | 13.2 Framing/ reframing  5.1 Information about health consequences  4.1 Instruction on how to perform the behaviour  13.2 Framing /reframing  6.1 Demonstration of the behaviour  7.1 Prompts/cues  2.2 Feedback on behaviour  2.3 Self-monitoring of behaviour  8.1 Behavioural practice/rehearsal  3.1 Social support (unspecified)  3.2 Social support (practical)  6.2 Social comparison | 33. Ensure PrEP information, training, education, and other communications directed at SHCPs are harmonised with the goals of the PrEP programme (i.e. explicit that PrEP is inclusive and relevant to all those with an identified need, not just MSM (13.2)  58. SHS could consider outsourcing educational sessions for SHCPs to CBOs with expertise on the specific sexual health cultures of and HIV risks affecting Black Africans, trans people, and cisgendered women (5.1)  59. SHS could ask CBO staff who have high levels of cultural competency in delivering sexual health promotion interventions to Black Africans, trans people, and cisgendered women to share their tailored vocabularies and co-produce a stock of key phrases to enable SHCPs to sensitively probe clients when taking a sexual/ drug history (4.1, 6.1, 7.1)  13. Review and update the questions asked as part of a sexual/drug history on a regular basis to ensure they reflect the epidemiological evidence and any emerging new trends or behaviours which appear to enhance the risk of HIV and cascade any changes to all staff (4.1)  34. Ensure SHCPs maintain their knowledge of the HIV risks among different groups, including MSM, and skills in conducting culturally sensitive clinical risk assessments (e.g. ongoing CPD, clinical supervision) (5.1, 2.2, 2.3, 8.1)  14. Adopt a protocoled approach to PrEP that includes advice (e.g. clear statements and nuanced examples) regarding the eligibility criteria (4.1, 13.2)  16. Ensure a range of peer-support systems are in place (e.g. real-time/email support, team meetings, ‘phone a friend’, clinical network arrangements) to assist SHCPs in making complex eligibility decisions (12.2, 3.1, 3.2, 6.2) | 33. Keep (no discussion).  58. Keep but modify (discussed) – not outsourcing, ‘SHCPs may benefit from close working with…’ Open the door to partnership work but recognising the expertise may already be present in services. Merge with 59. *  59. Keep (discussed) – merge with 58. Specific example under the main heading of working with CBOs / cultural sensitivity. *  13. Kill (discussed) – standard practice.  34. Keep but modify (no discussion) - avoid specifying MSM if talking about all PrEP beneficiaries. This links in with 5 and 6 and would work well combined. Could also include something along the lines of ‘think PrEP’. *  14. Keep (no discussion). *  16. Keep (no discussion) – could be linked to 5. In the longlist this recommendation was merged with the one re: peer-support systems to assist SHCPs in making complex decisions on medical suitability for PrEP (4 pages down). * |
|  |  | SHCPs find it easy to identify PrEP candidates based on risk of HIV acquisition because they can discuss and confirm or decide on (in)eligibility with a colleague | *“Personally, I've only seen one person who I discussed with a consultant, and they fell into* *that category. They were intending to travel as a sex tourist, and the number of partners that they were indicating was really quite significantly high. So, I deferred to someone else, to make that final decision.”* (SHCP) | Environmental context and resources  Social influences | Environmental restructuring  Enablement | 12.2 Restructure the social environment  3.1 Social support (unspecified)  3.2 Social support (practical)  6.2 Social comparison | 54. Facilitate and sustain a respectful team-oriented culture that values and promotes open exchange of ideas and perspectives and allows opportunities to foster good working relationships (e.g. huddles, team meetings) (12.2)  16. Ensure a range of peer-support systems are in place (e.g. real-time/email support, team meetings, ‘phone a friend’, clinical network arrangements) to assist SHCPs in making complex eligibility decisions (12.2, 3.1, 3.2, 6.2) | 54. Kill (no discussion) – not specific to PrEP. |
|  |  | SHCPs find it easy to identify PrEP candidates based on risk of HIV acquisition because they are trained in communication skills and experienced in taking comprehensive sexual/drug histories as part of their routine practice | *“I: How did you get those skills?*  *R: Through years of experience probably. I make it so matter of fact as if it’s conversation and I think a lot of my colleagues do the same. That, do you know what, I’ve got all these questions to ask. Feel free to say no, and in all the years I’ve been doing it I could count on my one hand how many people would say* *I’m not answering that, no. And then if they were to say that, I would be, and can I ask you why? No, so I just, I make it as part of conversation to be honest and I do say that, you know, I have had a lot of years’ experience, so I think it’s just the way I am.”* (SHCP) | Skills  Behavioural regulation  Professional role and identity | Training  Modelling  Enablement | 4.1 Instruction on how to perform the behaviour  6.1 Demonstration of the behaviour  8.1 Behavioural practice/rehearsal  2.2 Feedback on behaviour  2.3 Self-monitoring of behaviour  12.2 Restructuring the social environment | 45. Use a multi-method approach (e.g. online modules, shadowing, clinical supervision) to train SHCPs in communication skills (4.1), including observable best practice examples (e.g. video, in-person) (6.1) and time to practice (8.1), receive feedback (2.2), and reflect on their skills (2.3)  46. Allow opportunities for shared learning and peer reflections to enhance the communication skills of new and/or less experienced SHCPs, for example, through the introduction of a mentoring scheme (12.2) | 45. Kill (discussed) – generic and already covered.  46. Kill (no discussion) – applies generically rather than to PrEP specifically. |
|  |  | SHCPs find it easy to identify PrEP candidates based on risk of HIV acquisition because supporting documents and the IT system guide what issues to cover when taking a sexual/drug history | *“All these questions [to identify risk of HIV acquisition] are on our NaSH computer system anyway, but we also do have them all on our [paper-based] proforma.”* (SHCP) | Environmental context and resources  Behavioural regulation | Training  Enablement | 4.1 Instruction on how to perform the behaviour  7.1 Prompts/cues  2.3 Self-monitoring of behaviour | 17. Create paper-based or electronic protocols and proformas that provide SHCPs with clear guidance on the key questions to ask when taking a sexual/drug history (4.1, 7.1)  18. Introduce interactive ‘pop-up’ messages within the IT system that alert SHCPs to clients who appear eligible for PrEP based on the answers to key questions asked when taking a sexual/ drug history (7.1) and require them to confirm they have discussed PrEP with the client (2.3) | 17. Kill but ensure in stage 1 (discussed) – agreed guidance is needed, especially for HCPs less familiar with PrEP / in primary care. Very important to extending the reach of PrEP.  18. Kill (discussed) – pop-up messages don’t work / are hated. |
| SHCPs decide on the safety of prescribing and medical suitability for PrEP | SHCPs find it difficult to decide on the safety of prescribing and medical suitability for PrEP because the novelty of PrEP meant they were worried about making the ‘wrong’ decision, with anxieties amplified by the idea that PrEP prescribing needed to be consultant-led | SHCPs find it easy to decide on the safety of prescribing and medical suitability for PrEP because they are familiar with the drug through prescribing PEP and delivering HIV care, perceive it to be well-tolerated, and are used to prescribing for new indications | *“There was a level of anxiety about what the contraindications to PrEP were, what the renal toxicity might be. And it’s one thing for those clinicians, particularly doctors, who have cared for HIV patients for a long time, so have used the PrEP drugs for other reasons, but a lot of the clinical team weren’t necessarily involved in HIV care. So, they had a level of concern about what the toxicity issues might be, making an adequate assessment of any underlying conditions, worrying about bone health and other medications et cetera. And I have to say I think that still exists in quite a lot of settings.”* (SHCP) | Knowledge  Beliefs about consequences  Beliefs about capabilities  Skills | Education  Persuasion  Training | 5.1 Information about health consequences  13.2 Framing/ reframing  4.1 Instruction on how to perform the behaviour  6.1 Demonstration of the behaviour  8.1 Behavioural practice/rehearsal  8.7 Graded tasks  2.2 Feedback on behaviour  2.3 Self-monitoring of behaviour | 35. Ensure SHCPs are educated about PrEP via a comprehensive and ongoing training package that covers HIV testing, the HIV window period, and risk of antiretroviral resistance, common side-effects and their typically transient nature, the likelihood of toxic effects and role of monitoring to prevent long-term issues, and contraindications (5.1)  36. Demystify PrEP and build SHCPs confidence by presenting PrEP as a drug that can be prescribed by any qualified prescriber or supplied via agreed protocols (e.g. PGD) within SHS settings (13.2)  19. Produce national guidelines to promote and instruct SHCPs on safe prescribing of and medical suitability for PrEP, review and update the guidelines to reflect new information and lessons learned over time, and cascade any changes to all staff (5.1, 4.1)  20. Develop a tool that yields ‘okay to prescribe’ (green), ‘discuss with a colleague’ (amber), and ‘do not prescribe’ (red) outcomes (i.e. based on the traffic light system) to help SHCPs assess medical suitability for PrEP (5.1, 4.1)  37. Inform SHCPs that they can easily access up-to-date and evidence-based online information on interactions between PrEP and other drugs (e.g. www.hiv-druginteractions.org) (4.1)  38. Train SHCPs on how to conduct adequate assessments of any underlying health conditions and interpret the results of new tests required to establish medical suitability for PrEP (4.1, 6.1), share example cases for SHCPs to discuss and work through (8.1, 8.7), provide feedback (2.2), and allow opportunities for ongoing reflections on skill acquisition (2.3) | 35. Keep (no discussion)  36. Keep (no discussion) – links to 33, could combine.  19. Keep but modify (did not discuss) – consider cutting the last bit about cascading any changes to all staff (communication issue separate to the need for national guidelines).  20. Kill (no discussion) – decisions need to be nuanced and engage both patient and HCW to ensure equity. Also not sure that PrEP prescribing is complicated enough to warrant such a tool.  37. Keep (discussed) – very specific example, could be included in a box of ‘top tips’.  38. Keep (no discussion). |
|  | SHCPs find it difficult to decide on the safety of prescribing and medical suitability for PrEP because of conflicting advice and mixed messages from senior colleagues | SHCPs find it easy to decide on the safety of prescribing and medical suitability for PrEP because of formal and informal opportunities for peer support, for example, to seek advice, check and share decision-making, and discuss more medically complex cases, at local-, regional-, and national-level | “*We’re learning as we go along…and that’s maybe why we’re getting conflicting opinions from experts…we are coming across things we maybe didn’t anticipate early on. And so, it's useful to share that information…and if you're at a meeting like that, you tend to get a consensus opinion, there's maybe three or four experts there. And each expert will give their opinion on whatever case is presented, so you can reach some sort of consensus there. So, I think that’s quite helpful, for the more challenging cases.”* (SHCP) | Environmental context and resources | Environmental restructuring  Enablement | 12.2 Restructuring the social environment  3.1 Social support (unspecified)  3.2 Social support (practical)  6.2 Social comparison | 7. Use national infrastructure to facilitate discussion among senior HIV clinicians and reach a consensus on best practice for a range of scenarios to promote consistency in decisions on the safety of prescribing and medical suitability for PrEP (12.2, 3.1. 3.2)  Ensure there are formal and informal peer-support systems at local-, regional-, and national-level (e.g. real-time/email support, team meetings, ‘phone a friend’, clinical network arrangements) to assist SHCPs in making complex decisions on medical suitability for PrEP (12.2, 3.1, 3.2, 6.2) | 7. Keep but modify (no discussion) – change to ‘senior clinicians’ instead of ‘senior HIV clinicians’.  Keep (discussed) – was merged with recommendation 16 for the longlist hence no number. So SHCPs need peer-support systems to assist them in making complex decisions on eligibility and medical suitability for PrEP. |
|  | SHCPs find it difficult to decide on the safety of prescribing and medical suitability for PrEP because they do PrEP on a sporadic rather than regular basis | SHCPs find it easy to decide on the safety of prescribing and medical suitability for PrEP because they learn from exposure and ‘on the job’ experience | *“M1: How do you assess whether someone is straightforward or not, has that changed as time goes on?*  *F4: Yeah, yeah, definitely, it's definitely changing.*  *I1: How has this changed?*  *F4: From case discussions, from advice from others, from mistakes, you know. So it's definitely a learning process. Experience, really, and the more exposure to it has definitely changed the way that I think, and assess people. And what the follow-up is as well.”* (SHCPs) | Environmental context and resources  Memory, attention, and decision processes  Behavioural regulation | Environmental restructuring  Enablement  Training | 12.2 Restructure the social environment  3.1 Social support (unspecified)  3.2 Social support (practical)  4.1 Instruction on how to perform the behaviour | 21. Within SHS with high numbers of clients starting PrEP and initiating PrEP via specific clinics, rotate SHCPs to allow for an intense learning period that will enable them to become more decisive, confident, and expert about making decisions on the safety of and medical suitability for PrEP over time (12.2)  39. Acknowledge that all SHCPs are ‘learning on the job’ and restructure the social environment to allow opportunities for shared learning and peer reflections (12.2, 3.1, 3.2)  19. Produce national guidelines to promote and instruct SHCPs on safe prescribing of and medical suitability for PrEP, review and update the guidelines to reflect new information and lessons learned over time, and cascade any changes to all SHCPs (4.1) | 21. Kill (no discussion) – can train staff effectively without rotation.  39. Kill (no discussion) – not PrEP specific.  19. Keep but modify (did not discuss) – consider cutting the last bit about cascading any changes to all staff (communication issue separate to the need for national guidelines). |
|  | SHCPs find it difficult to decide on the safety of prescribing and medical suitability for PrEP because of limited opportunities to take up education and training (e.g. no slack in the system to free up staff, few clients on PrEP) | SHCPs find it easy to decide on the safety of prescribing and medical suitability for PrEP because of formal and informal training and learning opportunities at local-, regional-, and national-level | *“It’s taken much longer for some of the nursing team to develop the confidence because there isn’t that same opportunity for them to sit in a clinic and see a whole series of patients with the same problem. You know, certainly in the beginning it wasn’t even one a week, it was one every several weeks would come in and the you just have to take the opportunity that the right person was there. And that’s meant that some of the nursing team have felt really very exposed and it’s a real challenge for them.”* (SHCP) | Environmental context and resources  Knowledge | Enablement  Environmental restructuring  Education  Training | 3.2 Social support (practical)  12.2 Restructuring the social environment  5.1 Information about health consequences  5.3 Information about social and environmental consequences  9.1 Credible source  4.1 Instructions on how to perform the behaviour  6.1 Demonstration of the behaviour | 1. Those that fund SHS should provide the resource required to match the innovation (i.e. increase the budget according to predicted PrEP demand to ensure adequate capacity for effective implementation and scale-up) in the initial months of national rollout (3.2) A business case produced by senior HIV clinicians that outlines the health benefits of PrEP (5.1) and potential future savings of PrEP implementation within the healthcare system (i.e. more cost-effective than spending on HIV treatment) (5.3) could be helpful in this regard (9.1)  40. Offer a range of formal and informal opportunities for SHCPs to train and learn about safe prescribing of and medical suitability for PrEP, for example, at local- (e.g. journal clubs, team meetings, study days, shadowing), regional- (e.g. clinical network arrangements), and national-level (e.g. shared learning events) (12.2)  41. National coordinated PrEP training should include inter-disciplinary online PrEP learning resources for SHCPs which can be broken down into short modules on specific topics (e.g. covering safe prescribing of and medical suitability for PrEP) and spread out over a period of time (5.1, 4.1). These could be aligned with CPD for many job roles (12.2)  42. Introduce a shadowing scheme across different SHSs to enable SHCPs from SHS with few PrEP users to become familiar with PrEP processes, including ensuring safe prescribing of and medical suitability for PrEP (12.2, 6.1) | 1. Keep but modify (discussed) – replace ‘innovation’ with ‘clinical activity / costs of the programme’. Delete ‘produced by senior HIV clinicians’.  40. Keep (discussed) – range of learning opportunities provides equity for staff who like to learn in different ways. Training should be similarly accessible whether in a city / rural setting. Norming the values of PrEP – this is what we do, we offer PrEP, we’ve bought into it. Vital part of training and support for a national programme. *  41. Keep (discussed) – we are overemphasising toxicity throughout, however, online learning resources could be useful as part of a comprehensive training package. Passive compared to active. Merge with 42 and 43 as examples of a good training programme. *  42. Keep (discussed) – interactive and supportive training. Merge with 41 and 43 as examples of a good training programme. |
|  |  | SHCPs find it easy to decide on the safety of prescribing and medical suitability for PrEP because booked PrEP appointments provide the opportunity to review electronic patient records in advance and seek help, if necessary | *“What I tend to do is I review the clinic before I do it. So, like I reviewed the clinic yesterday for a guy, and I knew there was one guy who had got some particular medical conditions and I wanted to discuss that with the HIV consultants before I saw him, and I've done that, and I now have a plan and I know to go ahead and safely do things.”* (SHCP) | Environmental context and resources  Behavioural regulation | Environmental restructuring  Training  Education  Modelling  Enablement | 12.1 Restructure the physical environment  12.2 Restructure the social environment  5.1 Information about health consequences  5.3 Information about social and environmental consequences  6.1 Demonstration of the behaviour  2.2 Feedback on behaviour  2.3 Self-monitoring of behaviour | 22. In the initial stages of PrEP roll-out, consider implementing PrEP through booked appointments (12.1) to allow SHCPs the opportunity to review electronic patient records and seek any necessary help (e.g. research an issue, consult a colleague) before starting clients on PrEP (12.2)  Promote the advantages of high-quality clinical record keeping (5.1, 5.3), share best practice examples that meet the standards set out by the SHS and/or relevant professional bodies (6.1), and appraise and encourage SHCPs to reflect on their skills of recording episodes of care (2.2, 2.3) | 22. Keep but modify (no discussion) – change wording to ‘…to allow sufficient time for SHCPs the opportunity who may be unfamiliar with PrEP to review electronic patient records…’. |
| SHCPs communicate ineligibility for PrEP | SHCPs find it difficult to communicate ineligibility for PrEP because they feel under pressure from patients to prescribe and lack the knowledge, skills, and experience to convey the risk-benefit of PrEP for the individual | SHCPs find it easy to communicate ineligibility for PrEP because they avoid using the eligibility criteria terminology and instead focus the discussion on the risk-benefit of PrEP for the individual | *“I think the terminology is a real problem. It should be, is this the best thing for you, does your HIV risk outweigh the potential risk of rental toxicity in you. And that isn't conveyed, with the terminology – you're eligible, you're not eligible. And I think that terminology makes patients really angry. And I think that is probably one of the biggest problems, is telling people, you're not eligible. I think that people really don't like being told that. Whereas, if they're told, well actually, your HIV risk is actually this, statistically…or is this amount, but actually, your risk of renal damage if we give you this drug, is this amount. So actually, on balance, for you, I would recommend that you use condoms...”* (SHCP) | Environmental restructuring  Knowledge  Skills  Professional role and identity | Environmental restructuring  Education  Training  Persuasion  Modelling | 12.2 Restructure the social environment  13.2 Framing/ reframing  5.1 Information about health consequences  4.1 Instructions on how to perform the behaviour  7.1 Prompts/cues  6.1 Demonstration of the behaviour  8.1 Behavioural practice/rehearsal  8.7 Graded tasks  2.2 Feedback on behaviour | Throughout PrEP provision and promotion (e.g. during SHS- and CBO staff-client interactions, in national patient information booklets, on SHS, CBO, and HIV/ PrEP activists’ websites and social media, in marketing campaigns) avoid using the term ‘eligibility criteria’ and instead adopt ‘needs-based’ terminology that explicitly conveys PrEP decisions as a function of the individual risk-benefit of PrEP for each client (12.2, 13.2)  43. Ensure SHCPs are educated, trained, and appraised in their skills in discussing the risks and benefits of PrEP (e.g. through online modules, peer support, clinical supervision), for example, by giving information on PrEP health consequences (5.1), producing a ‘how to’ script for common PrEP scenarios based on the lessons learned of SHCPs with general medicine expertise (4.1, 7.1), and providing opportunities to shadow (6.1), practice with (8.1, 8.7), and receive feedback (2.2) from more experienced SHCPs | Keep (not discussed) – did not include in longlist of recommendations due to being so close to recommendation 67 hence no number. Merge with 67.  43. Keep (discussed) – Claudia likes the wording. Merge with 41 and 42 as examples of a good training programme. Relates to 33? * |
|  |  | SHCPs find it easy to communicate ineligibility for PrEP because they view the interaction as a teachable moment and can frame ineligibility in a positive light | *“R: That can be the tricky thing to say to people – you're not eligible.*  *I: How do those conversations go?*  *R: So sometimes I feel like it's quite useful, because you can revisit all the other prevention strategies with them, and you can kind of congratulate them and say, you know, I think you're managing your risk really well yourself. And usually, I think if you take time to explore that side of things with people, then it goes fine.”* (SHCP) | Professional role and identity  Social influences | Education  Persuasion  Enablement | 5.1 Information about health consequences  10.4 Social reward  13.2 Framing/ reframing  1.2 Problem solving  3.1 Social support | 70. SHCPs should reassure clients that they are at low risk for HIV by educating them (e.g. verbally, directing to reputable websites) on the facts of HIV transmission and effectiveness of alternative sexual health promotion methods (5.1)  71. SHCPs could congratulate clients on their safer sex practices (10.4) and suggest that they consider their not needing PrEP as a positive outcome (i.e. they are already sufficiently protected against HIV) (13.2)  72. SHCPs should explore the root cause(s) of HIV-related anxieties among clients who do not have an identified need for PrEP and work with them to problem solve solutions (1.2) | 70. Keep (discussed) – can be merged with 62.  71. Kill (discussed) – generic that you acknowledge when people are doing well / do not require an intervention and reinforce the ‘good’ behaviour.  72. Keep (no discussion). |
|  |  | SHCPs find it easy to communicate ineligibility because they can suggest self-sourcing PrEP online and still offer PrEP monitoring within the healthcare system | *“I say, if you really, you really want to put medicine in you, and we don't really need you to, you know. And they say, well I really do, and then you say, well you can buy it yourself, and we talk about self-buying. Because we can, like the NHS can, like, check the side-effects, and look after people who are self-buying themselves, you know. So, I can still refer into [clinic], if they're gonna buy it themselves.”* (SHCP) | Knowledge  Environmental context and resources | Enablement  Environmental restructuring | 3.1 Social support  12.1 Restructure the physical environment | 74. SHCPs need to be aware of the option to self-source PrEP and could consider directing clients who do not meet the eligibility criteria but would still like to access PrEP to reputable online sources of information about where to buy PrEP (e.g. provision of national patient information booklets, signpost to iwantPrEPnow.co.uk) (3.1)  23. Expand the remit of SHS to include providing care for those self-sourcing PrEP (12.1) and ensure that clients are aware of when and how they can access PrEP monitoring (e.g. SHCPs provide information verbally, hand out location-specific leaflets or wallet-sized inserts, signpost to websites) (3.1) | 74. Keep (no discussion) –  overlap with 66 – merge?  23. Keep (no discussion). |
|  |  | SHCPs find it easy to communicate ineligibility for PrEP because they can make explicit reference to the eligibility criteria within which they are permitted to prescribe as a fall back and means of justification | *“I would say to them, you know, this is a government led thing, they're paying for it, and this is the reason why there is certain criteria… this is what has been said, it's not what I'm saying, it's what somebody else has written down and we've got to follow the guidelines. So I think when people realise that, then they're okay with it. It's not that you're making that decision, so I would sit with the guidelines and go through them one by one with like the criteria, and go through them and say you don't fit any of them.”* (SHCP) | Memory, attention, and decision making  Environmental context and resources  Professional role and identity | Training  Enablement  Environmental restructuring  Persuasion | 4.1 Instruction on how to perform the behaviour  13.2 Framing /reframing  12.1 Restructure the physical environment  9.1 Credible source | 14. Adopt a protocoled approach to PrEP that includes advice (e.g. clear statements and nuanced examples) regarding the eligibility criteria (4.1, 13.2)  15. Advise SHCPs to keep a copy of the PrEP protocol handy (12.1) and make explicit reference to it when communicating their decision not to prescribe PrEP to clients (9.1), being careful not to shut down more wholistic conversations | 15. Kill (no discussion) – service detail, not specific to PrEP. |
| SHCPs explain the different PrEP regimens | SHCPs find it difficult to explain the different PrEP regimens because of the complexity of and unfamiliarity with on-demand dosing (e.g. when to start, stopping rules for different scenarios) | SHCPs find it easy to explain the different PrEP regimens because national patient information booklets that detail the key points about the various ways to take PrEP and provide diagrams showing how to follow on-demand dosing serve as an aide memoire and can be used to structure the conversation | **Example 1**  *“I don’t know how good I would be if they were saying so I’m going to have sex on a Saturday and then I’m going to have sex on a Thursday, when do I actually start and stop it, you know. So, it’s case-by-case and I probably still need to refresh my memory a little bit and read up a bit on that still if I was doing that because most of the people are just taking it every day.”* (SHCP)  **Example 2**  *“You go through the i-Base PrEP Scotland leaflet with them, because they’ve got some nice information and diagrams just to explain the difference between event-based and daily.”* (SHCP) | Knowledge  Memory, attention, and decision processes  Environmental context and resources | Education  Enablement | 4.1 Instruction on how to perform a behaviour  7.1 Prompts/cues  2.7 Feedback on behaviour | 44. Use a multi-method approach to educate SHCPs about on-demand dosing (4.1) and assist them during consultations (7.1). For example:  Develop a range of resources (e.g. brief fact sheet, PrEP provider pocket guide, national patient information booklets) with clear written instructions and diagrams that depict how to take PrEP on-demand, including examples of when to start and stop for various scenarios, which can be used to educate SHCPs (4.1) and assist them during consultations (7.1). Such resources should ideally be co-produced by a range of diverse organisations and the communities who will use them)  Provide SHCPs with laminated copies of the on-demand dosing diagrams that they can pin to their wall as a quick reminder of how to take PrEP on-demand (4.1, 7.1)  Record a short video or soundbite that explains on-demand dosing for different scenarios that SHCPs may watch or listen to at a future date (4.1)  Include an online or paper-based quiz with questions about on-demand dosing as part of SHCPs PrEP training and ongoing CPD and ensure that there is sufficient time or a named person to contact to discuss the answers after, if necessary (2.7) | 44. Keep (discussed) – both SHCPs and PrEP users struggle with event-based / intermittent dosing. It needs to be explained better so people can take PrEP appropriately, since most HIV diagnoses among people on PrEP are in those taking it event-based / intermittently. These practical suggestions may work well as specific examples of training. |
| *PrEP uptake* | | | | | | | | |
| Potential PrEP users accurately report their sexual/drug history | Potential PrEP users find it difficult to accurately report their sexual/drug history because otherwise they will not get access to PrEP (i.e. they do not meet the eligibility criteria) | Potential PrEP users find it easy to accurately report their sexual/drug history because the availability of PrEP allows for a more worthwhile discussion on HIV risks and thus prompts more open and honest disclosures | **Example 1**  *“I’ve had friends that lied about it and said, yeah, I’ve had four unprotected partners in the last three weeks, just so they could get on it. And if I hadn’t had the right sort of qualification, I guess, to get on it, I would have probably lied as well. I would have said, I had six partners last week, if that was what it would have taken to be able to get on it.”* (*PrEP user*)  **Example 2**  *“I just know that certainly there are more guys prepared to tell you and be more open and honest about the kind of sex that they’re having that they weren’t going to have been before because they weren’t going to get anything to make that better. Whereas now they can chat to me about it and know that I’m able to give them PrEP if nothing else.”* (SHCP) | Knowledge  Beliefs about consequences  Social influences | Education  Persuasion  Training  Enablement | 5.1 Information about health consequences  13.2 Framing/ reframing  4.1 Instruction on how to perform the behaviour  6.1 Demonstration of the behaviour  8.1 Behavioural practice/rehearsal  8.7 Graded tasks  2.2 Feedback on behaviour  9.1 Credible source | 67. Ensure PrEP information and communications (e.g. SHS- and CBO staff-client interactions, national patient information booklets, SHS, CBO, and HIV/PrEP activists’ websites and social media, marketing campaigns) avoid using the term ‘eligibility criteria’ and instead adopt ‘needs-based’ terminology that explicitly conveys the risks and benefits of PrEP and thus the rationale against its blanket supply (5.1, 13.2)  32. Ensure SHCPs are educated (5.1), trained (4.1, 6.1, 8.1, 8.7), and appraised in their skills (2.2) in explaining the risk-benefit of PrEP and mandate this activity in a formal protocol (4.1, 5.1)  68. SHCPs should explain to clients the importance of reporting an accurate sexual/ drug history to ensure they receive the most appropriate care to their individual needs (5.1, 9.1)  69. SHCPs should actively promote PrEP to clients as one of several sexual health promotion methods (5.1) and emphasise their own and other experts and credible sources’ support for it (e.g. government, public health agencies, CBO staff) (9.1) so clients feel comfortable to disclose their HIV risks | 67. Keep but modify (not discussed) – consider deleting ‘and thus the rationale against its blanket supply’.  69. Keep but modify (discussed) – remove the section after 9.1. |
|  | Potential PrEP users find it difficult to accurately report their sexual/drug history because they expect to be judged by or pick up on judgement from a SHCP about their lifestyle, sexual norms, and relational dynamics | Potential PrEP users find it easy to accurately report their sexual/drug history because SHCPs present as friendly and approachable, engage in active listening, and conduct culturally sensitive clinical risk assessments | **Example 1**  *“R: There are still some stigmas around partner numbers, the types of sex that people are having, the number of partners at a given time, group sex, a lot of things, drug use, alcohol use, that become almost quite finger waggy.*  *I: What does finger waggy mean?*  *R: As in disapproving, and there’s a moral judgement that comes with clinical risk assessment, and patients can pick up on that, and they pick up on it really, really quickly, and that just wrecks a patient’s consultation and you’re probably never going to get it back.”* (SHCP)  **Example 2**  *“It's a question of just listening a little bit more. Not having a dismissive attitude. I think everybody likes to be listened to. And it's really important, when people, even if they are speaking with an accent, to try and listen, and try to understand where they are coming from. Which really makes a huge difference, to the way people will behave after that.”* (CBO staff working with Black African communities) | Environmental context and resources  Professional role and identity  Social influences | Environmental restructuring  Training  Enablement  Persuasion | 12.2 Restructure the social environment  5.3 Information about social and environmental consequences  4.1 Instruction on how to perform the behaviour  6.1 Demonstration of the behaviour  7.1 Prompts/cues  5.1 Information about health consequences  9.1 Credible source | 55. Facilitate and maintain (e.g. via training, huddles, clinical supervision, reflective practice) a warm, welcoming, and friendly atmosphere wherein SHCPs communicate with clients in a non-judgemental manner, using active listening and inclusive, sex- and PrEP-positive, and destigmatising language to establish trust and ensure an open dialogue (12.2, 5.3)  59. SHS could ask CBO staff who have high levels of cultural competency in delivering sexual health promotion interventions to communities affected by HIV to share their tailored vocabularies and co-produce a stock of key phrases to enable SHCPs to sensitively probe clients when taking a sexual/drug history (4.1, 6.1, 7.1)  56. SHS should assure potential PrEP users that the SHS is a welcoming, safe, and non-judgemental space through co-produced (e.g. with CBO staff, community representatives) culturally appropriate literature (e.g. posters, national patient information booklets) in SHS waiting areas and consultation rooms and other settings (e.g. at CBOs, GP) and online information (e.g. via SHS websites and social media) (12.2)  68. SHCPs should explain to clients the importance of reporting an accurate sexual/ drug history to ensure they receive the most appropriate care to their individual needs and reassure them that SHS operate to a particularly high standard of confidentiality (5.1, 5.3, 9.1) | 55. Keep but modify (not discussed) – though this recommendation received 2 kills and 1 keep, coinvestigators were clear in the meeting about ensuring we retain elements such as providing a non-judgemental, stigma-free, and supportive environment. A recommendation of this nature has to feature somewhere.  56. Keep but modify (discussed) – quite a meaty recommendation. Want to ensure we retain the bit on SHS being a welcoming, safe, and non-judgemental (stigma-free, supportive environment) but separate out the key message about the importance of having co-produced culturally appropriate literature available in a range of settings (i.e. not just in SHS), including online. *  68. Kill (no discussion) – SHS offer anonymity rather than more confidentiality, but not sure it is helpful to suggest SHS are better providers in that respect. It is extremely desirable to encourage users to trust other services such as GP to meet sexual health care needs including PrEP. |
|  | Potential PrEP users find it difficult to accurately report their sexual/drug history because they have very low levels of sexual health and HIV literacy and struggle to talk about their sexuality, sexual health, and HIV prevention needs (e.g. because of cultural stigmas attached to sex and a history of being underserved in sex education) |  | *“Most of the time people are not going to confess, because for us, it’s taboo to talk about sex. We grew up when our parents are telling you that it’s a bad thing, so literally you don’t discuss sex, whatever happens.”* (CBO staff working with Black African communities) | Environmental context and resources  Knowledge | Environmental restructuring  Education | 12.2 Restructure the social environment  5.1 Information about health consequences | 8. Governments should make age-appropriate and comprehensive relationships and sex education compulsory for children and young people at all levels of schooling, with fact-oriented and non-judgemental content that addresses the sexual health, social, and cultural needs of LGBTQ+ and Black African communities (12.2), incorporating issues such as PrEP as HIV prevention (5.1)  75. CBO staff and other HCPs, such as GPs, must address cultural stigmas and normalise talking about sexuality, sexual health, and HIV prevention needs by engaging clients and the wider communities that they serve in topics of this nature (e.g. via discussions in everyday contexts / routine consultations, interactional workshops at diverse community venues, outreach work) (12.2) | 8. Kill (discussed) – contentious issue, don’t like use of word ‘compulsory’, less crucial than other recommendations, not specific to PrEP.  75. Keep but modify (discussed) – important to include and like the wording. Need to differentiate non-clinical staff such as health improvement to do this to make it practical. Limited scope to get GPs to do this, but probably mean primary care in general. Relates to the need for sexual health / PrEP training to be relevant to all settings. * |
| Potential PrEP users take up PrEP | Potential PrEP users find it difficult to take up PrEP because SHCPs push PrEP too much, or not enough, and provide personal perspectives rather than expert opinion | Potential PrEP users find it easy to take up PrEP because SHCPs discuss PrEP via a balanced narrative that reinforces individual choice and is supportive of taking time to consider whether PrEP is right for them | **Example 1**  *“He was kind of telling me about all the good things about PrEP, but I wasn’t...I don’t know. I didn’t want to buy it, if this is a phrase, because he was almost saying that it’s the best thing ever, because he was using it, he was using it and he told me that. So, I don’t know, I kind of stopped using the [clinic].”* (PrEP user)  **Example 2**  *“I think her words were, have you thought about PrEP? She sort of prompted it, prompted the conversation but didn’t push it and then I continued the conversation. So it wasn’t a you’ve had unprotected sex, you should take PrEP, it was more of a have you considered this drug? Do you know about the benefits it could bring to you? It was, she prompted it, but didn’t push it on me in any way.”* (PrEP user) | Environmental context and resources  Professional role and identity  Skills  Social influences | Environmental restructuring  Education  Persuasion  Enablement  Training  Modelling | 12.2 Restructure the social environment  5.3 Information about social and environmental consequences  5.1 Information about health consequences  2.3 Self-monitoring of behaviour  2.4 Self-monitoring of the outcome(s) of behaviour  5.3 Information about social and environmental consequences  5.6 Information about emotional consequences  16.3 Vicarious consequences  9.2 Pros and cons  3.1 Social support  4.1 Instruction on how to perform the behaviour  6.1 Demonstration of the behaviour | 47. Educate SHCPs on the importance of the SHS and/or relevant professional bodies’ ethical principles, policies, and code of conduct (5.3, 5.1) and engage them in regular reflective practice (2.3, 2.4) to ensure that they maintain appropriate professional boundaries (12.2)  76. SHCPs should draw on research evidence and what they know about other clients’ decision-making and experiences to inform clients of the health, social, and emotional benefits of PrEP (5.1, 5.3, 5.6, 16.3) but also stress that PrEP is a choice and discuss the pros and cons of taking up PrEP compared to not taking up PrEP with respect to clients’ individual interests (9.2)  77. SHCPs should avoid pressurising clients to take up PrEP by suggesting that they take time to consider their decision (12.2) and directing them to alternative reputable information sources (e.g. provision of national patient information booklet, signposting to SHS, CBO, and HIV/PrEP activists’ websites and social media) (3.1)  48. Provide informal learning opportunities (e.g. during team meetings) for SHCPs to share successful conversation approaches for pitching PrEP to clients (4.1, 6.1) | 47. Kill (no discussion) – applies generically rather than specifically to PrEP.  76. Keep (no discussion).  77. Kill (no discussion).  48. Kill (no discussion). |
|  | Potential PrEP users find it difficult to take up PrEP because they are dubious about the validity of PrEP (i.e. distrust in PrEP) | Potential PrEP users find it easy to take up PrEP because they are aware of the research evidence and/or have heard statements of support for the efficacy and safety of PrEP by an expert source (e.g. SHCP, CBO staff) | *“I was very sceptic about it at first because I wasn’t sure exactly how it works, but then once I read through research about it online and also the clinician gave me a booklet explaining what it was. So, once I started getting it, I was more comfortable with it and I found it is safe.”* (PrEP user) | Knowledge  Professional role and identity | Education  Persuasion | 5.1 Information about health consequences  9.1 Credible source | 78. PrEP information and communications (e.g. SHCP-/ CBO staff-client interactions, national patient information booklets, SHS, CBO, and HIV/PrEP websites and social media, posters in SHS and CBO settings, marketing campaigns) should  provide an accessible, scientific explanation of what PrEP does (i.e. how it works inside the body) and describe PrEP efficacy and safety with reference to key research and ‘real world’ studies and regional or national HIV incidence data (5.1, 9.1) | 78. Keep but modify (discussed) – add in something about the information being presented in culturally appropriate language, co-produced. People from the diverse communities who may benefit from PrEP have to see themselves represented. Links into 56. |
|  | Potential PrEP users find it difficult to take up PrEP because they do not want to take a (daily) pill for HIV prevention | Potential PrEP users find it easy to take up PrEP because of its flexibility | **Example 1**  *“Taking a drug for anything is not something that I particularly want to do… and it took me a little bit of time to get round in my head to take a drug preventatively.”* (PrEP user)  **Example 2**  *“You can always stop taking it if you decided you weren’t comfortable with the side effects if there were any, or if a test came back that your kidney function wasn’t great, or as a result of taking PrEP it was worse, you just stop taking it.”* (PrEP user) | Beliefs about consequences  Environmental context and resources  Memory, attention, and decision processes | Enablement  Education  Persuasion | 13.2 Framing/ reframing  5.1 Information about health consequences | 79. All sectors involved in PrEP should consider a range of approaches (e.g. via SHCP-/CBO-client interactions, SHS, CBO, and HIV/PrEP activists’ websites and social media, national patient information booklets, marketing campaigns) to: normalise PrEP by drawing parallels to the use of daily preventive medicine in other areas of health (e.g. contraceptive pill to protect against pregnancy, blood thinners to reduce the risk of heart attack and stroke) (13.2); and educate potential PrEP users on the flexibility of PrEP by informing them of the idea of ‘seasons of risk’ (i.e. unlikely to be on PrEP forever, can start and stop as circumstances dictate) and the various dosing options (i.e. can opt for less intensive on-demand dosing, if appropriate) (5.1, 13.2) | 79. Keep (no discussion). |
|  | Potential PrEP users find it difficult to take up PrEP because of the perceived negative health and social consequences of PrEP (e.g. side effects, changes to their own and other people’ sexual behaviours, STIs, PrEP stigma, cost of PrEP and burden on the healthcare system) | Potential PrEP users find it easy to take up PrEP because of the perceived positive health, emotional, and social consequences of PrEP (e.g. taking responsibility for their own and other people’s sexual health, contributing to ending the HIV epidemic, cost of PrEP compared to HIV treatment, alleviating personal HIV fear, sexual freedom and enjoyment, social acceptability) | **Example 1**  *“I think it encourages perhaps having more sex, having more one-night stands or more promiscuous sex and doing so without being protected. And we both know that there’s a whole host of other diseases out there, other than HIV, which are just as dangerous. Even though they can be cured they can have complications on your health also.”* (Declined PrEP when offered)  **Example 2**  *“If I had unprotected sex, there would be a period of six weeks to three months where I was not entirely sure whether or not I had contracted HIV. And that's not a very comfortable feeling to live with day-to-day. And it's just not a very healthy way to think about sex and think about intimacy. I think that probably was the biggest driver, was just this wanting to have an intimacy that wasn't connected with fear. And I think PrEP provides that.”* (PrEP user) | Beliefs about consequences | Education  Persuasion | 5.1 Information about health consequences  5..3 Information about social and environmental consequences  5.6 Information about emotional consequences  9.1 Credible source  1.2 Problem solving  13.2 Framing/ reframing  6.2 Social comparison | 9. Coinciding with PrEP roll-out, public health agencies, health authorities, and others with a remit for sexual health promotion should commission a mass media/social marketing campaign aimed at reaching those who may benefit from PrEP. This could be fronted by culturally appropriate opinion leaders and would aim to share news of recent advancements within the HIV field (e.g. U=U, PrEP) and inform about the economic and wider benefits and value of PrEP for the healthcare system, communities, and individuals (5.1, 5.3, 5.6, 9.1)  61. SHCPs and CBO staff should inform clients (e.g. verbally, provision of national patient information booklets, signposting to SHS, CBO, and HIV/PrEP activists’ websites and social media, talks at CBO events, positive PrEP user testimonials) of the health, cost/financial, social, and emotional impacts of PrEP (5.1, 5.3, 5.6, 9.1) to enable them to make informed choices about whether or not to take up PrEP  81. SHCPs should educate clients about the potential side-effects of PrEP and their typically transient nature (5.1), share management strategies for the most common side-effects (1.2), and reassure against concerns about longer-term toxic effects by drawing attention to the tests undertaken at three-month reviews (5.1)  62. SHCPs and CBO staff should encourage PrEP users to continue using condoms alongside PrEP by framing PrEP as an additional rather than alternative HIV prevention method (13.2)  80. Normalise PrEP by informing clients and potential PrEP users not already accessing SHS of PrEP uptake data for the SHS, region, and/or country (e.g. via a marketing campaign, verbally by SHCP and CBO staff, peer education) (6.2) | 9. Keep but modify (no discussion) – needs to be done as really important to not assume that community links (apps, venues etc.) will be the only way people learn about PrEP. But there were political reasons why PrEP was not advertised widely in 2017, so at what stage? Has to be reconciled with concerns about capacity within the clinic and recognise that large scale public health campaigns are inherently political and guided by politics. Also has to be combined with specific targeted approach for non-white and non-MSM. *  61. Kill (no discussion) – cannot think how a service might be delivered without, as we do for all interventions.  81. Keep (no discussion) – though this recommendation received 2 kills and 1 keep, concerns about side effects were frequently mentioned as a reason why participants were wary of taking up PrEP.  62. Keep but modify (discussed) – encourage thinking beyond PrEP or nothing. Remove emphasis on condoms, add combination prevention, and tailoring to individual. Merge with 70.  80. Keep – link to other recommendations about normalisation. |
|  |  | Potential PrEP users find it easy to take up PrEP because of the support, encouragement, and positive PrEP experiences of important others (e.g. peers, partners, friends, family) | *“I guess half my influence was from my friend and the other was from online reviews. And the kind of advice that my friend gave me was I might feel sort of dizzy or sickly for the first one or two weeks of taking PrEP, but that’s just my body getting used to it. And since they’ve started taking PrEP they’ve become a lot more... they felt a lot more comfortable about having unprotected sex.”* (PrEP user) | Social influences  Beliefs about consequences | Enablement  Education  Persuasion | 3.1 Social support (unspecified)  1.2 Problem solving  5.1 Information about health consequences  5.3 Information about social and environmental consequences  5.6 Information about emotional consequences  9.1 Credible source  4.1 Instruction on how to perform the behaviour  1.4 Action planning  8.1 Behavioural practice/rehearsal | 63. SHCPs and CBO staff should encourage clients to discuss PrEP with important others (3.1), ask them to identify any potential barriers, and select strategies to overcome these (1.2)    64. SHCPs and CBO staff should persuade PrEP users to share PrEP information and talk about their own PrEP experiences with important others by informing them of the important health, social, and emotional benefits of doing so (e.g. increase awareness and uptake of PrEP, reduce PrEP-related stigma) (5.1, 5.3, 5.6, 9.1)  65. SHCPs and CBO staff should support clients to have PrEP conversations by prompting detailed planning of what they want to convey (e.g. for PrEP users - the meaning and benefits of PrEP for them, how they overcame any issues), how (e.g. suggest key phrases to use) (4.1), when, and to whom (1.4) and facilitating a role-play exercise(s) as practice (8.1) | 63. Kill (discussed) – not specific to PrEP, discussions with important others encouraged for other heath interventions. Too basic.  64. Keep but modify (discussed) – remove ‘persuade’ and use ‘encourage where appropriate’ instead.  65. Kill (no discussion) – not practical and unknown effectiveness. |
| Potential PrEP users choose their preferred PrEP regimen | Potential PrEP users find it difficult to choose their preferred PrEP regimen because SHCPs only offer the option of daily PrEP or provide a strong personal opinion indicating a preference for daily PrEP | Potential PrEP users find it easy to choose their preferred PrEP regimen because SHCPs explain the options in an unbiased manner and engage in a shared decision-making process | *“He said, I prefer people just to take it every day, because then they're covered. He said, what happens is, people forget to take it when they're taking it event-based and they don't take it in time, and then they're actually not protected by it properly. He said, taking it every day means you're completely protected whether you meet people or you don't meet people.”* (PrEP user) | Memory, attention, and decision processes  Environmental context and resources  Professional role and identity  Skills  Social influences | Environmental restructuring  Education  Persuasion  Enablement  Training  Modelling | 12.2 Restructure the social environment  5.3 Information about social and environmental consequences  5.1 Information about health consequences  9.2 Pros and cons  6.1 Demonstration of the behaviour | 57. Educate and persuade SHCPs to ensure a non-paternalistic service environment (i.e. respectful of clients’ right to choice and characterised by shared decision-making) is provided and sustained over time (12.2, 5.3, 5.1)  82. SHCPs should inform clients of their options for how to take PrEP by way of a balanced narrative (5.1) and then jointly, with each individual client, facilitate a decisional balance weighing up the pros and cons per option, taking into account lifestyle and/or the availability of evidence to support it (i.e. dependent on gender and whether oral, anal, or vaginal/frontal sex) (9.2)  49. Identify SHCPs who are particularly skilled at supporting clients to choose their preferred PrEP regimen and ask them to demonstrate their approach (e.g. via video example, role-/real-play exercises, shadowing) for other SHCPs to imitate (6.1) | 57. Kill (no discussion).  82. Keep (no discussion).  49. Kill (discussed) – not really practical as it’s difficult to identify who these people are. |
|  |  | Potential PrEP users find it easy to choose their preferred PrEP regimen because there is a wealth of information on PrEP dosing options online | *“Everybody accesses information on the net but PrEP’s more extreme because that’s where it started, with people investigating online. So to a greater extent than, for example, women taking the contraceptive pill, people taking PrEP will seek advice on what to do from non-medical sources.”* (SHCP) | Environmental context and resources | Enablement | 3.1 Social support  9.1 Credible source | 66. SHCPs and CBO staff could direct clients to reputable online sources of information on the various ways to take PrEP (e.g. SHS, CBO, and HIV/PrEP activists’ websites and social media) (3.1, 9.1) in addition to the information they provide (e.g. verbally, via provision of national patient information booklet) | Keep (no discussion). |
| Potential PrEP users get their first PrEP prescription | Potential PrEP users find it difficult to get their first PrEP prescription because of the necessary pre-assessment tests | Potential PrEP users find it easy to get their first PrEP prescription because they opportunistically see a SHCP who makes a clinical judgement to prescribe PrEP before the pre-assessment test results are back | **Example 1**  *“We ideally need to rule out the fact that you’re not seroconverting and that all your kidney functions are all working. So we need to wait for those test results to come back before you get prescribed.”* (SHCP)  **Example 2**  *“If it’s a point of care test, then we’ll most likely give it on the day. Or if somebody has a significant risk and won’t reduce their risk for a week, or can’t rather than won’t, then you have to make a clinical decision about whether we just get it started and take the risks that are attached.”* (SHCP) | Environmental context and resources | Environmental restructuring | 12.1 Restructure the physical environment | 24. In line with WHO guidelines, PrEP providers should move to routine use of point of care rapid HIV tests and starting clients on PrEP on the same day that they present to SHS, with the exception of special circumstances (e.g. exposure to HIV in the last 72 hours, signs/symptoms of acute HIV infection, known renal issues) and so long as they agree to be contacted and return to see a SHCP if any of the baseline test results require action, confirmation, or treatment (12.1) | 24. Kill (discussed) – difficult practicalities in some settings for POC testing, hard to implement. Would mean prescribing PrEP before receiving results of renal tests as standard. Might help people who struggle to attend sexual health services as only one visit required. Suitably rapid / 24-hour turnaround of 4^th^ gen HIV tests may be more desirable. WHO guidelines may actually mean POC test is better than not testing (which does happen in some settings). |
|  | Potential PrEP users find it difficult to get their first PrEP prescription because PrEP is dispensed offsite and there are restrictions on which pharmacies can supply it | Potential PrEP users find it easy to get their first PrEP prescription because PrEP is dispensed onsite | **Example 1**  *“Initially we had to give them a hospital prescription to pick up their PrEP and because we’re in the community hospital, they had to pick up the hospital prescription from [organisation], which is in [place], it’s not the easiest place to get to, if you don’t have your own transport.”* (SHCP)  **Example 2**  *“She sort of went into another room and then came back with all the boxes. And I was like, oh, wow, that’s it...I remember being a bit surprised. Not in a negative way, just in a, oh, great sort of way.”* (PrEP user) | Environmental context and resources | Environmental restructuring | 12.2 Restructure the social environment  12.5 Adding objects to the environment  12.1 Restructure the physical environment | 25. Designate a qualified person within the SHS to be responsible for establishing a PrEP supply chain (12.2) and maintaining agreed stock levels (12.5) to enable SHCPs to dispense PrEP to clients during their PrEP appointment  10. Work with pharmacy leads to extend the role of community pharmacists to enable clients to obtain PrEP via a range of settings (12.1) | 25. Keep but modify (no discussion) – the important bit is having a one stop shop (i.e. prescribe and dispense not prescribe, go and queue at a pharmacy). Delete the bit re: a designated person (service detail, need a team) and make the other bit more explicit.  10. Keep (no discussion) –  promotes equity of access. |
